# Sex- and Age-Specific Associations of Blood Lead Levels with Child Growth and Haemoglobin in Bangladesh: A Nationally Representative Study

**DOI:** 10.64898/2026.11.07.26367484

**Authors:** Muhammad Mainuddin Patwary, Sadia Ashraf, Md Atikul Islam, Matthew H.E.M. Browning, Michelle L. Bell

**Author notes:** **Corresponding author:** Muhammad Mainuddin Patwary, Environment and Sustainability Research Initiative, Khulna, Bangladesh.

## Abstract

Evidence on sex- and age-specific associations between lead levels and childhood growth remain limited. We analyzed 10,324 children aged 12–59 months from the Bangladesh Multiple Indicator Cluster Survey 2025 using survey-weighted multivariable linear regression to examine sex- and age-specific association of blood lead level (BLL) with anthropometric measures and haemoglobin levels. The geometric mean BLL was 4.43 µg/dL, and 66.3% of children had concentrations above the Centers for Disease Control and Prevention (CDC) threshold (≥3.5 µg/dL). Higher BLL was associated with lower height-for-age z-score (HAZ) (β = −0.129, 95%CI: −0.182 to −0.076), weight-for-age z-score (WAZ) (β = −0.108, 95%CI: −0.156 to −0.060), and weight-for-height z-scores (WHZ) (β = −0.052, 95%CI: −0.101 to −0.002). Higher BLL was also associated with lower HAZ and WAZ among boys (HAZ: β = −0.114, 95%CI: −0.188 to −0.040; WAZ: β = −0.113, 95%CI: −0.179 to −0.046) and girls (HAZ: β = −0.150, 95%CI: −0.225 to −0.075; WAZ: β = −0.104, 95%CI: −0.171 to −0.036), while higher BLL was associated with lower WHZ among boys (β = −0.076, 95%CI: −0.144 to −0.007). Higher BLL was associated with lower HAZ (β = −0.266, 95%CI: −0.408 to −0.125) and WAZ (β = −0.206, 95% CI: −0.326 to −0.086) among boys aged 24–35 months, while among girls aged 36–59 months, corresponding estimates were −0.211 (95%CI: −0.310 to −0.113) and −0.106 (95%CI: −0.191 to −0.022). The findings suggest that longitudinal studies are needed to clarify the temporal and developmental patterns underlying these associations.

**Graphical Abstract:** 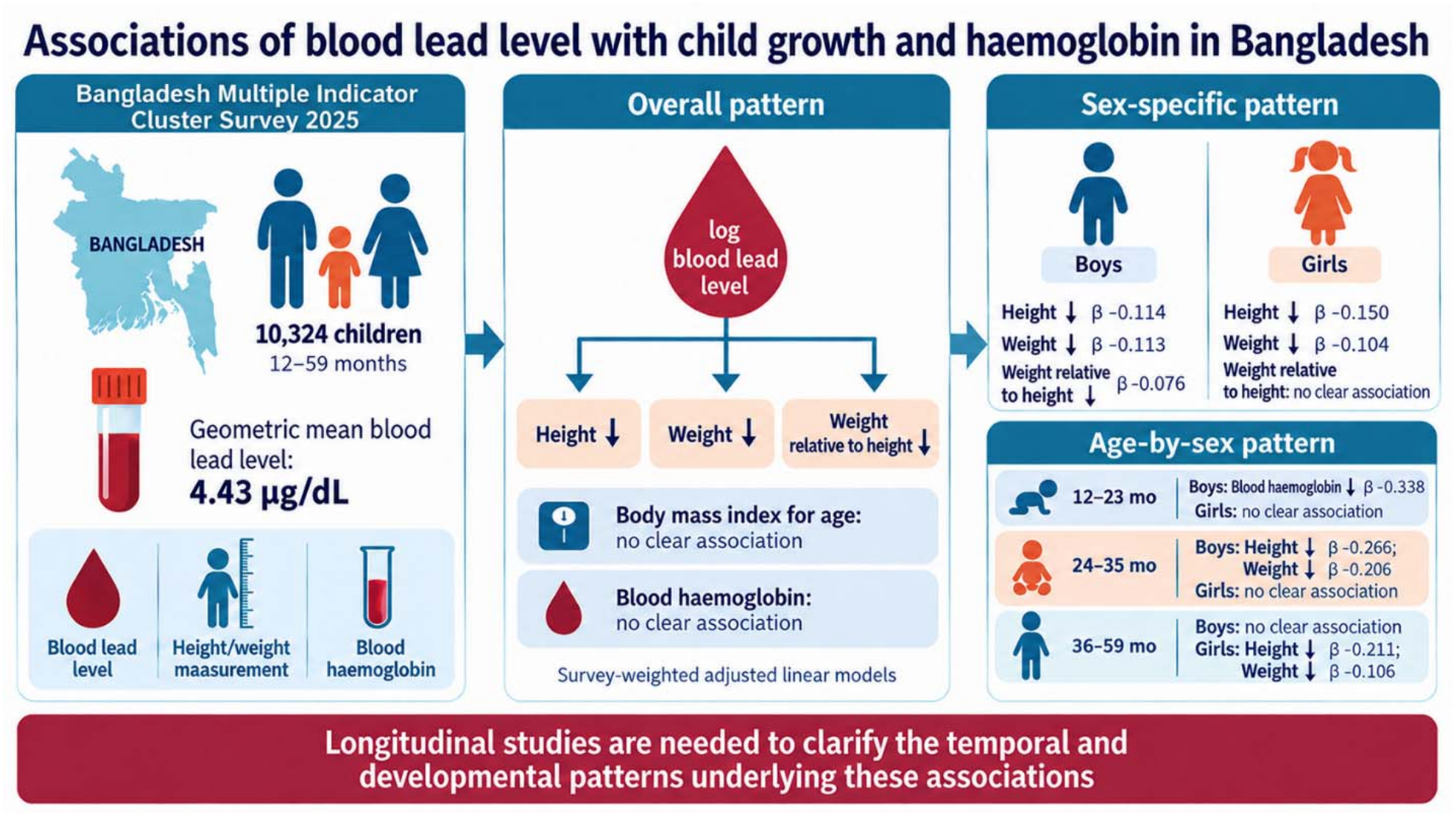

## 1. Introduction

Lead (Pb) is a widespread environmental toxicant that affects multiple organ systems and remains a global public health concern. Young children are particularly vulnerable because they absorb lead more efficiently than adults and are frequently exposed through hand-to-mouth behaviour and ingestion of contaminated dust, soil, food, or water (WHO, 2026). A joint UNICEF–Pure Earth assessment estimated that about 800 million children worldwide had blood lead concentrations of at least 5 µg/dL, which is equivalent to one in every three children (UNICEF and Pure Earth, 2020). This burden is disproportionately concentrated in low- and middle-income countries (LMICs) (Ericson et al., 2021), with South Asia accounting for a substantial share of the global burden (Roy and Das, 2026). The effects of lead may continue long after the exposure has occurred. A global modelling study estimated that lead exposure was associated with 765 million lost IQ points among children younger than five years in 2019 and with approximately 5.5 million adult cardiovascular deaths (Larsen and Sánchez-Triana, 2023).

Lead exposure has been associated with adverse neurological, haematological, renal, cardiovascular, and developmental outcomes, and no blood lead concentration has been established as being without harmful effects in children (WHO, 2026). Child growth is a clinically relevant marker of health and cumulative nutritional and environmental stress. However, the association between lead exposure and anthropometric status is not consistent across settings. In a study of young Chinese children, higher blood lead levels (BLL) were associated with lower height-for-age z-score (HAZ) and weight-for-age z-score (WAZ) (Zhou et al., 2020). In rural Bangladesh, concurrent BLL was associated with a higher odds of stunting among children aged 20–40 months, whereas cord-BLL was not associated with later stunting (Gleason et al., 2016). A separate study of children living in informal settlements in Bangladesh found association with stunting and underweight but not with wasting (Raihan et al., 2018). Findings from other populations have been mixed, with associations varying according to the timing and biomarker of exposure, the growth measure examined, and the age and sex of the child (Ahmadi et al., 2022). These inconsistencies suggest that a single overall association may conceal important developmental or sex-specific patterns.

Lead may influence haemoglobin through disruption of haem biosynthesis, including inhibition of enzymes in the pathway, and this effect may be compounded by iron deficiency, which can increase gastrointestinal lead absorption (Abelsohn and Sanborn, 2010; WHO, 2026). Nevertheless, epidemiological findings concerning blood lead and haemoglobin have been inconsistent, and relatively little evidence has examined whether this relationship differs across specific developmental periods (Irawati et al., 2022; Mitra et al., 2012). Lead may also affect growth through interference with endocrine and skeletal processes. For example, higher BLL have been associated with lower circulating insulin-like growth factor-1 in children, although the direction and clinical importance of this pathway remain uncertain (Fleisch et al., 2013; Roels et al., 1976). However, these mechanisms do not necessarily imply that associations should be uniform across childhood. Exposure patterns, nutritional status, rates of growth, physiological development, and susceptibility to toxicants can change substantially during the first five years of life (Abelsohn and Sanborn, 2010; Afeiche et al., 2012).

Sex and developmental age may therefore be important sources of heterogeneity in the association between lead exposure and child health. However, previous studies have not reached a clear conclusion about whether boys or girls are more affected by lead. In a study of 1,678 young children in China, higher blood lead was associated with lower HAZ and WAZ among boys but not girls, and the authors concluded that sex-specific patterns warranted further investigation (Zhou et al., 2020). Other studies have reported different patterns of evidence (Ashley-Martin et al., 2019; Malin Igra et al., 2021). These inconsistencies suggest that sex-specific susceptibility cannot be assumed to operate in a uniform direction. Age may be equally important. During the first five years of life, rapid changes in growth, behaviour, diet, and exposure pathways can affect lead exposure and nutritional needs, particularly because of frequent hand-to-mouth behaviour, greater gastrointestinal absorption, and the transition to complementary and family foods (Hauptman et al., 2017; WHO, 2026). For example, a study in Mexico suggested that infancy may be a sensitive period for lead exposure in relation to height at 48 months (Afeiche et al., 2012). However, evidence on whether lead exposure is associated differently with physical growth and haemoglobin across developmental age groups remains limited.

Despite the biological plausibility linking lead exposure to impaired growth and haemoglobin synthesis, evidence from LMICs, including Bangladesh, remains limited. Previous studies on lead and child growth have largely focused on rural communities, urban informal settlements or specific geographic areas, often with relatively small sample sizes and varying age ranges (Gardner et al., 2013; Gleason et al., 2016; Raihan et al., 2018). Findings have also been inconsistent, and few studies have examined blood lead, haemoglobin, and multiple anthropometric indicators within the same nationally representative population. Evidence on whether these associations differ by sex or developmental age is also limited.

The Bangladesh Multiple Indicator Cluster Survey (MICS) 2025 provides an opportunity to address these gaps. Covering all 64 districts, the survey collected blood lead, haemoglobin, and standardized anthropometric measurements among children aged 12–59 months and represents the first national household survey in Bangladesh to measure blood lead and other heavy metals at this scale (BBS and UNICEF, 2026). These data allow assessment of lead exposure and child health beyond the restricted populations examined previously. However, national prevalence estimates do not indicate whether blood lead is associated with specific growth or haemoglobin outcomes or whether these associations vary by age or sex. Therefore, this study aimed to examine the associations between blood lead concentration and child growth indicators, and haemoglobin among Bangladeshi children aged 12–59 months using the nationally representative MICS 2025 dataset. Specifically, we assessed the overall associations and evaluated whether they differed by sex and developmental age group.

## 2. Methods

### 2.1. Data source and participant selection

We used data from the Bangladesh MICS 2025, a nationally representative household survey conducted by the Bangladesh Bureau of Statistics (BBS) in collaboration with UNICEF as part of the global MICS programme. The survey was conducted from 24 February to 30 June 2025 using computer-assisted personal interviewing based on the 2022 Bangladesh Population and Housing Census. A multistage, stratified cluster sampling design was used to select households. The sampling framework comprised 67 sampling domains, including the 64 administrative districts and three separate city-corporation domains: Dhaka North, Dhaka South, and Chattogram. Within each domain, census Enumeration Areas (EAs) served as primary sampling units (PSUs) and were selected separately within urban and rural strata using probability-proportional-to-size sampling. Following household mapping and listing, 20 households were selected systematically from each sampled EA. Overall, 3,149 clusters (862 urban and 2,287 rural) and 62,980 households were selected across Bangladesh. The survey included household, women, and child questionnaires, along with a biomarker component for children aged 12–59 months and pregnant women. The present study included children aged 12–59 months with available measurements of BLL, haemoglobin, and anthropometric outcomes (BBS and UNICEF, 2026).

The Bangladesh MICS 2025 included 24,680 eligible children under five years of age. Of these, 18,913 children were aged 12–59 months and were included in the population eligible for the blood lead and other heavy-metal assessment. Under the blood collection procedure, 17,132 children were eligible for blood collection, of whom 10,634 yielded a valid laboratory result. Further, we excluded participants with invalid or out-of-range anthropometric measurements or haemoglobin levels, resulting in 10,488 participants. Children with missing covariate information were also excluded and the final analytical sample comprised 10,324 children aged 12–59 months. The participant selection process is shown in Figure S1.

### 2.2. Blood collection and biomarker assessment

Blood biomarkers were assessed as part of the Bangladesh MICS 2025 biomarker component. Children aged 12–59 months were eligible for blood collection. In households with eligible children, one child was selected for blood collection. However, when more than one child aged 12–59 months was present, selection was performed using the Kish grid incorporated into the computer-assisted interviewing system. Blood collection was conducted at the household level by trained phlebotomists using aseptic precautions and sterile, disposable needles and syringes. Approximately 2 mL of venous blood was collected into metal-free tubes and transported to the Immunobiology, Nutrition and Toxicology Laboratory at the International Centre for Diarrhoeal Disease Research, Bangladesh (icddr,b) in Dhaka for laboratory analysis. The biomarker component was implemented with technical support from the Institute of Epidemiology, Disease Control and Research (IEDCR) and icddr,b (BBS and UNICEF, 2026).

### 2.3. Blood lead exposure assessment

Blood lead concentration was determined in whole blood using inductively coupled plasma mass spectrometry (ICP-MS). Blood specimens were digested with concentrated nitric acid and analysed using calibration standards and internal standards. Blanks were used to assess background contamination and carryover, while three levels of ClinChek control materials were used for quality assurance of analytical accuracy and precision. Blood lead concentration was reported in μg/dL and was used as the biomarker of internal lead exposure (BBS and UNICEF, 2026). In the present study, blood lead concentration was analysed both continuously and categorically. Because of its right-skewed distribution, blood lead was natural-log transformed for continuous analyses. For categorical analyses, blood lead was classified as <3.5 μg/dL and ≥3.5 μg/dL, with <3.5 μg/dL as the reference category. The 3.5 μg/dL threshold corresponds to the CDC blood lead reference value for children (CDC, 2021).

### 2.4. Anthropometric and haemoglobin measurements

Anthropometric measurements were collected as part of the standard MICS under-five survey procedures. Children’s weight and height/length were measured by trained field teams following standardized procedures. WAZ, HAZ, weight-for-height z-scores (WHZ), and body mass index-for-age z-scores (ZBMI) were derived according to the WHO Child Growth Standards (BBS and UNICEF, 2026). In the present study, WAZ, HAZ, WHZ, and ZBMI were included as continuous measures of child growth and nutritional status.

Haemoglobin was measured from whole blood using the cyanmethemoglobin method. Whole blood was mixed with Drabkin’s solution for 30 minutes, followed by measurement of optical density at 540 nm using a UV–Vis spectrophotometer. Haemoglobin concentration was calculated using a three-point calibration curve prepared with cyanmethemoglobin standards, with commercial quality-control blood used to monitor measurement performance (WHO, 2011). In the present study, haemoglobin concentration (g/dL) was treated as a continuous outcome measure.

### 2.5. Covariates

Covariates were selected a priori based on previous literature (Ashley-Martin et al., 2019; Gardner et al., 2013; Gleason et al., 2016; Raihan et al., 2018; Zhou et al., 2020), together with the availability of relevant variables in the Bangladesh MICS 2025 dataset (BBS and UNICEF, 2026). The adjustment set included child-level characteristics, maternal characteristics, household socioeconomic and environmental characteristics, and potential lead-exposure-related factors. Child-level covariates included age (months), age group (12–23, 24–35, and 36–59 months) and sex (male/female). Maternal characteristics included maternal age (years) and educational attainment (pre-primary/no education, primary, lower secondary, secondary, higher secondary, and higher). Household socioeconomic status was measured using the MICS wealth index, a composite score derived by principal component analysis (PCA) from household ownership of durable assets, dwelling characteristics and access to basic services (Rutstein and Rojas, 2006). Households were ranked by their standardized wealth score and divided into five equal quintiles: poorest, poor, middle, rich and richest. Place of residence was categorized into urban and rural. Administrative division was included as an eight-category variable representing Bangladesh’s eight divisions. Household environmental characteristics included the main source of drinking water and the type of sanitation facility, each classified as improved, unimproved, or other/unspecified following the WHO/UNICEF Joint Monitoring Programme (JMP) definitions used in MICS (WHO, 2025). Improved drinking-water sources included piped water, tubewells/boreholes, protected dug wells and springs, rainwater, packaged water, and delivered water, whereas unprotected wells and springs and surface water were classified as unimproved. Improved sanitation facilities included flush or pour-flush toilets connected to a piped sewer system, septic tank or pit latrine, ventilated improved pit latrines, pit latrines with slab, and composting toilets, while flush toilets discharging to an open drain, pit latrines without slab, bucket or hanging toilets, and open defecation were classified as unimproved. Other responses were grouped as other/unspecified. Housing characteristics included interior wall condition (no paint, paint in poor condition, or paint in good condition) and floor material (natural/rudimentary or finished). Household smoking was defined based on whether smoking was reported during the preceding three months. Two additional binary variables were constructed from the MICS lead-exposure module. Presence of a lead-related activity near the household was coded as “yes” if any of six activities reported in the survey: vehicle-battery manufacture, vehicle-battery recycling by melting, vehicle-battery repair or disassembly, jewelery manufacture or recycling, e-waste recycling, or automobile repair; otherwise, it was coded as “no”. Similarly, lead-related occupational exposure among child caregiver was coded as “yes” if any of the same six activities was reported for such an individual during the preceding three months, and “no” otherwise (BBS and UNICEF, 2026).

### 2.6. Statistical analysis

Descriptive characteristics were summarized using means and standard deviations for continuous variables and survey-weighted percentages for categorical variables. BLLs w summarized using the geometric mean, median, and interquartile range because of their right-skewed distribution, and distributions were examined across sex and age groups. Spearman correlation coefficients among continuous predictors, BLL, and study outcomes conducted (Figure S3). The correlations were generally modest, supporting the inclusion of the predictors in multivariable models (Figure S3). Variance inflation factors (VIFs) were also examined to assess multicollinearity (VIF > 5) (Table S1).

Blood lead was primarily analyzed as a continuous exposure after natural-log transformation. Associations with HAZ, WAZ, WHZ, ZBMI, and haemoglobin concentration were estimated using survey-weighted multivariable linear regression, with β coefficients and 95% CIs reported. BLL was additionally categorized as <3.5 versus ≥3.5 µg/dL, with <3.5 µg/dL as the reference group. The 3.5 µg/dL cut-off was based on the CDC BLL reference value and was used as an analytical threshold rather than a level indicating absence of harm. Associations were subsequently examined separately by sex and age group to assess whether the relationship between BLL and the outcomes differed across developmental stages and between boys and girls. Age was categorized into three groups: 12–23, 24–35, and 36–59 months. Sex-specific models were fitted separately for boys and girls, while age-specific models were fitted separately within each age group. Additional analyses examined associations within combined age-by-sex groups to describe patterns across developmental stages and sex. Variables corresponding to the stratification factor were omitted from the corresponding stratified models; for example, sex was not included in sex-specific models and age was not included as a continuous covariate in age-specific models. Sensitivity analyses assessed robustness to alternative exposure specifications by modeling BLL on the log scale, with effect estimates interpreted per doubling of BLL. Potential non-linear exposure–response relationships were evaluated using restricted cubic spline models (knot = 4), with non-linearity assessed using Wald tests of the spline terms. These analyses were considered sensitivity analyses to assess whether the primary linear exposure specification adequately represented the observed associations.

All models incorporated sampling weights, strata, and primary sampling units using the survey package in R (version 4.3.1) to account for the complex sampling design of the Bangladesh MICS 2025. Models were adjusted for child sex, child age as a continuous variable (months), maternal age as a continuous variable (years), maternal education, household wealth, residence, administrative division, drinking-water source, sanitation, interior wall condition, floor type, household smoking, nearby lead-related activity, and lead-related occupational exposure among child caregiver. Sex- and age-specific models were additionally fitted for boys and girls and for children aged 12–23, 24–35, and 36–59 months; the corresponding stratification variable was omitted from each stratified model. All tests were two-sided with α = 0.05, and effect estimates were reported with 95% CIs.

### 2.7. Ethical approval

This study used anonymised secondary data from the Bangladesh Multiple Indicator Cluster Survey (MICS) 2025, conducted by the Bangladesh Bureau of Statistics with UNICEF support. The survey was approved by the icddr,b Ethical Review Committee in November 2024 (PR-24113). Written informed consent was obtained from mothers or legal guardians before children’s blood collection. Data were accessed through the UNICEF MICS platform in accordance with applicable data-use agreements. As this analysis involved only de-identified data, no additional ethical approval was required.

## 3. Results

### 3.1. Characteristics of the study population

Table 1 summarizes the characteristics of the study population. The analysis included 10,324 children aged 12–59 months from the Bangladesh MICS 2025 biomarker sample. The mean age was 37.1 months, and boys comprised the majority (52.57%) of the sample. Children aged 36–59 months represented the largest age group (54.81%). The mean maternal age was 28.1 years. Most mothers had secondary (27.96%) or lower secondary education (27.24%). The majority of children lived in rural households (70.64%). The largest regional shares were observed in Dhaka (24.07%) and Chattogram (23.43%). The distribution across household wealth quintiles was relatively balanced, although the poorest households represented the largest group (21.91%) and the richest represented 16.48%. Regarding housing conditions, most (53.46%) children lived in households with no paint on interior walls, windows or doors. Nearly half of households had natural or rudimentary flooring (46.20%). Almost all households reported an improved drinking-water source (99.22%), while 90.52% had an improved sanitation facility. Household smoking was reported for 51.83% of children. Lead-related exposure characteristics were common in the study population. More than half of the children lived near a lead-related activity (52.00%), while 12.48% had a caregiver engaged in lead-related work.

**Table 1.** Characteristics of the study populations (N = 10,324).

| Characteristic | N | Weighted estimate |
|---|---|---|
| **Child characteristics** |  |  |
| ***Sex*** |  |  |
| Boys | 5,416 | 52.57% |
| Girls | 4,908 | 47.43% |
| ***Age, months*** | 10,324 | 37.1 ± 0.2 |
| ***Age group*** |  |  |
| 12–23 months | 1,976 | 19.05% |
| 24–35 months | 2,654 | 26.14% |
| 36–59 months | 5,694 | 54.81% |
| **Maternal characteristics** |  |  |
| ***Maternal age, years*** | 10,324 | 28.1 ± 0.1 |
| ***Maternal education*** |  |  |
| Pre-primary or none | 649 | 6.28% |
| Primary | 1,982 | 19.45% |
| Lower secondary | 2,805 | 27.24% |
| Secondary | 2,883 | 27.96% |
| Higher secondary | 1,153 | 10.77% |
| Higher | 852 | 8.29% |
| **Household characteristics** |  |  |
| ***Residence*** |  |  |
| Urban | 2,576 | 29.36% |
| Rural | 7,748 | 70.64% |
| ***Region*** |  |  |
| Barishal | 1,052 | 5.14% |
| Chattogram | 2,083 | 23.43% |
| Dhaka | 1,739 | 24.07% |
| Khulna | 1,575 | 10.39% |
| Mymensingh | 618 | 7.94% |
| Rajshahi | 1,125 | 11.47% |
| Rangpur | 1,467 | 11.15% |
| Sylhet | 665 | 6.40% |
| ***Household wealth index*** |  |  |
| Poorest | 2,716 | 21.91% |
| Poor | 2,436 | 21.52% |
| Middle | 2,089 | 20.27% |
| Rich | 1,814 | 19.83% |
| Richest | 1,269 | 16.48% |
| **Housing conditions** |  |  |
| ***Interior wall condition*** |  |  |
| No paint on walls, windows or doors | 6,085 | 53.46% |
| Paint in poor condition | 1,212 | 13.16% |
| Paint in good condition | 3,027 | 33.38% |
| ***Floor type*** |  |  |
| Natural/rudimentary | 5,414 | 46.20% |
| Finished | 4,877 | 53.34% |
| Other/unspecified | 33 | 0.46% |
| ***Drinking-water source*** |  |  |
| Unimproved | 59 | 0.45% |
| Improved | 10,223 | 99.22% |
| Other/unspecified | 42 | 0.33% |
| ***Sanitation facility*** |  |  |
| Unimproved | 1,001 | 9.29% |
| Improved | 9,305 | 90.52% |
| Other/unspecified | 18 | 0.20% |
| ***Household smoking*** |  |  |
| No | 4,947 | 48.17% |
| Yes | 5,377 | 51.83% |
| **Lead-related exposure characteristics** |  |  |
| ***Any lead-related activity nearby*** |  |  |
| No | 5,177 | 48.00% |
| Yes | 5,147 | 52.00% |
| ***Caregiver engaged in lead-related work*** |  |  |
| No | 9,176 | 87.52% |
| Yes | 1,148 | 12.48% |

### 3.2. Blood lead level (BLL)

The distribution of BLL illustrated in Table 2 & Figure 1. Overall, the geometric mean (GM) was 4.43 µg/dL, with a median of 4.32 µg/dL (IQR, 3.14 µg/dL) and arithmetic mean of 5.22 µg/dL (SD, 3.80) (Table 2). The highest estimated BLL value was observed in Dhaka (Figure S2). BLL were higher among boys than girls (GM: 4.54 vs 4.30 µg/dL; P < 0.001) and varied across age groups, with the highest GM was observed among children aged 12–23 months (4.69 µg/dL; P < 0.001) (Table 2 & Figure 1). Consistently, 66.26% of children had BLL ≥3.5 µg/dL, with a higher proportion among boys than girls (68.48% vs 63.80%; P < 0.001) and among children aged 12–23 months compared with older age groups (70.68%, 66.09%, and 64.80%, respectively; P < 0.001). Within each sex, age differences were not significant among boys (P = 0.257), whereas both continuous concentrations and the proportion ≥3.5 µg/dL declined significantly with age among girls (P < 0.001) (Table 2 & Table S2).

**Table 2.**
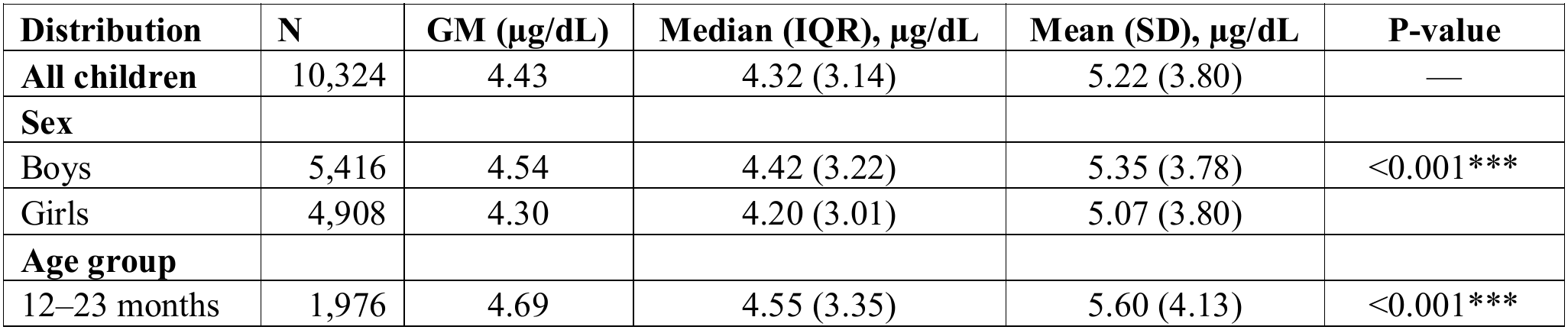
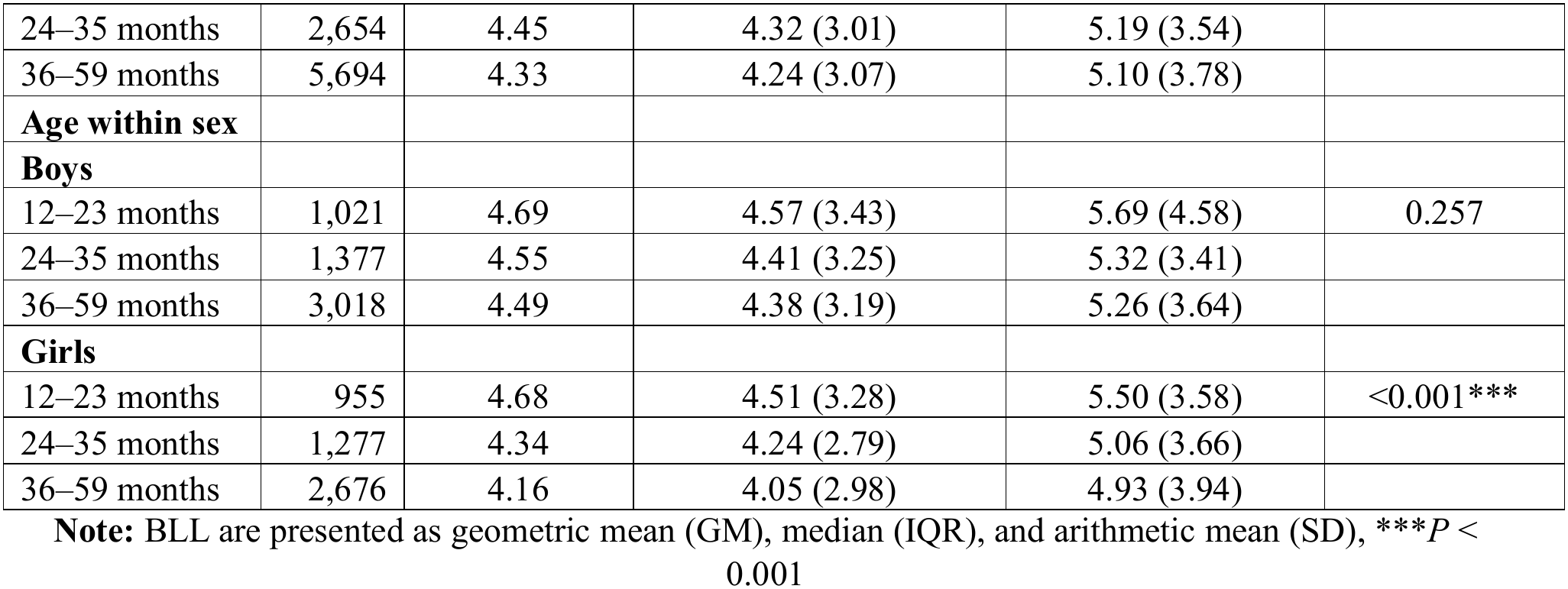
Distribution of BLL by sex and age group.

**Figure 1.**
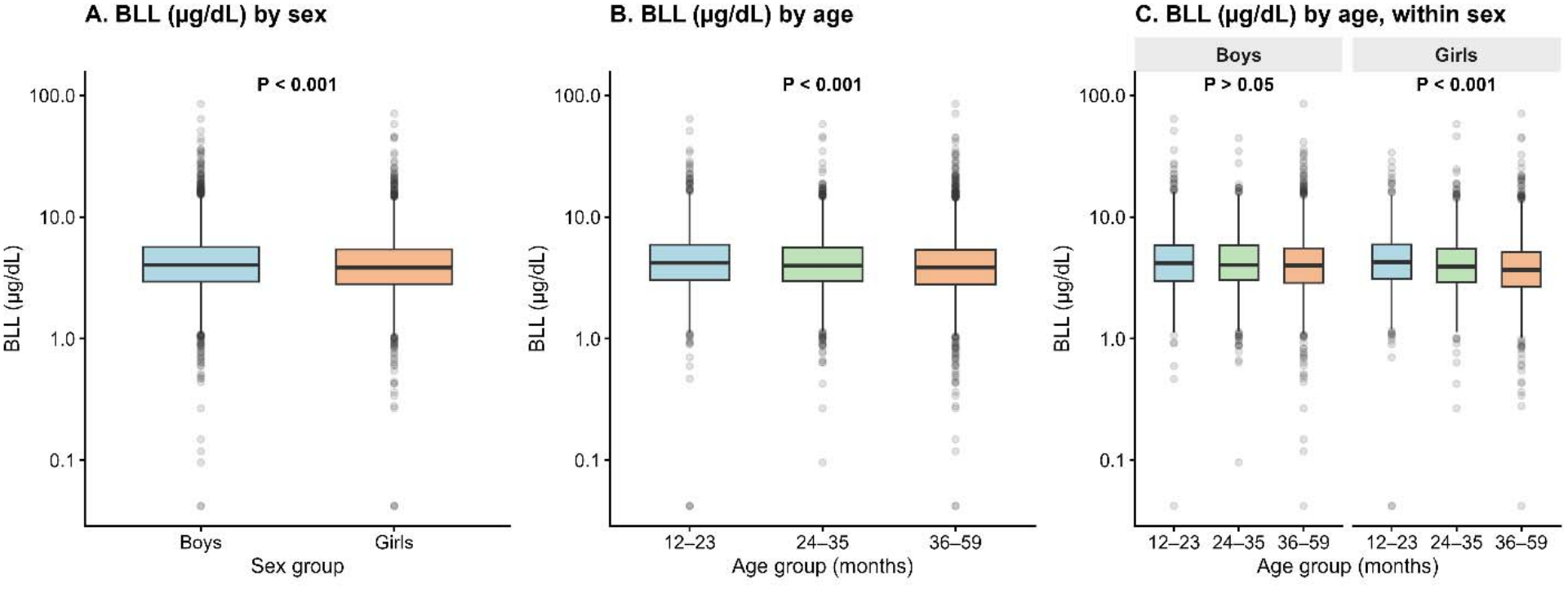
Boxplot showing the distribution of BLL (µg/dL) by sex and age group.

### 3.3. Distribution of growth and haemoglobin measures

Table 3 presents the distribution of child growth and haemoglobin measures. The overall mean of HAZ, WAZ, WHZ and ZBMI were −1.31, −1.38, −0.96 and −0.82, respectively, while mean haemoglobin concentration was 10.99 g/dL. Sex differences were less pronounced for HAZ, WAZ and ZBMI. WHZ was lower among boys than girls, with mean values of −0.99 and −0.92, respectively. Mean haemoglobin was also lower among boys than girls (10.95 vs 11.03 g/dL). Growth and haemoglobin measures showed variation across age groups. WAZ, WHZ, ZBMI and haemoglobin differed significantly across the three age groups, with the lowest mean haemoglobin observed among children aged 12–23 months (10.36 g/dL) and the highest among those aged 36–59 months (11.23 g/dL). Within sex, age-related differences were evident for WHZ, ZBMI and haemoglobin in both boys and girls, and for WAZ in girls only (P < 0.001); the age difference in WAZ was not significant among boys. Children aged 12–23 months had higher mean WHZ and ZBMI than older children in both sexes, and higher WAZ among girls, while haemoglobin increased progressively with age.

**Table 3.** Distribution of child growth and haemoglobin measures by sex and age group.

| **Distribution** | **N** | **HAZ** | **WAZ** | **WHZ** | **ZBMI** | **Hb** |
|---|---|---|---|---|---|---|
| **All children** | 10,324 | −1.31 (1.14) | −1.38 (1.03) | −0.96 (1.05) | −0.82 (1.06) | 10.99 (1.16) |
| **Sex** |  |  |  |  |  |  |
| Boys | 5,416 | −1.31 (1.16) | −1.38 (1.03) | −0.99 (1.07)** | −0.84 (1.07) | 10.95 (1.18)** |
| Girls | 4,908 | −1.32 (1.12) | −1.38 (1.03) | −0.92 (1.03) | −0.81 (1.06) | 11.03 (1.13) |
| **Age group** |  |  |  |  |  |  |
| 12–23 months | 1,976 | −1.30 (1.31) | −1.25 (1.12)*** | −0.85 (1.14)*** | −0.65 (1.17)*** | 10.36 (1.28)*** |
| 24–35 months | 2,654 | −1.36 (1.15) | −1.35 (1.05) | −0.88 (1.06) | −0.74 (1.08) | 10.93 (1.14) |
| 36–59 months | 5,694 | −1.30 (1.07) | −1.45 (0.98) | −1.03 (1.01) | −0.93 (1.00) | 11.23 (1.03) |
| **Age within sex** |  |  |  |  |  |  |
| **Boys** |  |  |  |  |  |  |
| 12–23 months | 1,021 | −1.33 (1.35) | −1.30 (1.13) | −0.90 (1.13)** | −0.69 (1.16)*** | 10.25 (1.34)*** |
| 24–35 months | 1,377 | −1.37 (1.18) | −1.37 (1.04) | −0.94 (1.07) | −0.77 (1.07) | 10.88 (1.13) |
| 36–59 months | 3,018 | −1.27 (1.07) | −1.42 (0.98) | −1.05 (1.04) | −0.91 (1.03) | 11.21 (1.02) |
| **Girls** |  |  |  |  |  |  |
| 12–23 months | 955 | −1.27 (1.26) | −1.20 (1.10)*** | −0.79 (1.14)*** | −0.59 (1.17)*** | 10.48 (1.20)*** |
| 24–35 months | 1,277 | −1.36 (1.13) | −1.34 (1.05) | −0.83 (1.04) | −0.71 (1.09) | 10.98 (1.14) |
| 36–59 months | 2,676 | −1.32 (1.07) | −1.47 (0.98) | −1.01 (0.97) | −0.94 (0.97) | 11.25 (1.03) |

Results expressed as mean (±SD); \**P* < 0.05, \*\**P* < 0.01, \*\*\**P* < 0.001; HAZ: Height-for-Age Z-score; WAZ: Weight-for-Age Z-score; WHZ: Weight-for-Height Z-score; ZBMI: Body Mass Index-for-Age Z-score; Hb: Haemoglobin.

### 3.4. Associations between BLL and child growth indicators and haemoglobin

#### 3.4.1. Overall associations

Figure 2 and Table S3 shows the results of overall associations of survey-weighted multivariable models. As shown in the model, higher continuous log-transformed BLL was associated with lower HAZ (β = −0.129, 95%CI: −0.182 to −0.076, P < 0.001), WAZ (β = −0.108, 95%CI: −0.156 to −0.060, P < 0.001), and WHZ (β = −0.052, 95%CI: −0.101 to −0.002, P < 0.05). However, no evidence of an association was found with ZBMI (P > 0.05) or haemoglobin concentration (P > 0.05). Similar patterns were observed when BLL was categorized by a threshold of 3.5 µg/dL. Compared with children with BLL below 3.5 µg/dL, those with BLL of 3.5 µg/dL or higher had lower HAZ (β = −0.087, 95% CI: −0.144 to −0.029, P < 0.01), WAZ (β = −0.084, 95% CI: −0.136 to −0.032, P < 0.01) and WHZ (β = −0.055, 95% CI: −0.109 to −0.001, P < 0.05). No significant associations were observed for ZBMI or haemoglobin (p > 0.05) (Figure 2 and Table S3). Sensitivity analysis using log_2_-transformed BLL (µg/dL) yielded similar patterns of associations, supporting the robustness of the primary findings (Table S4).

**Figure 2.**
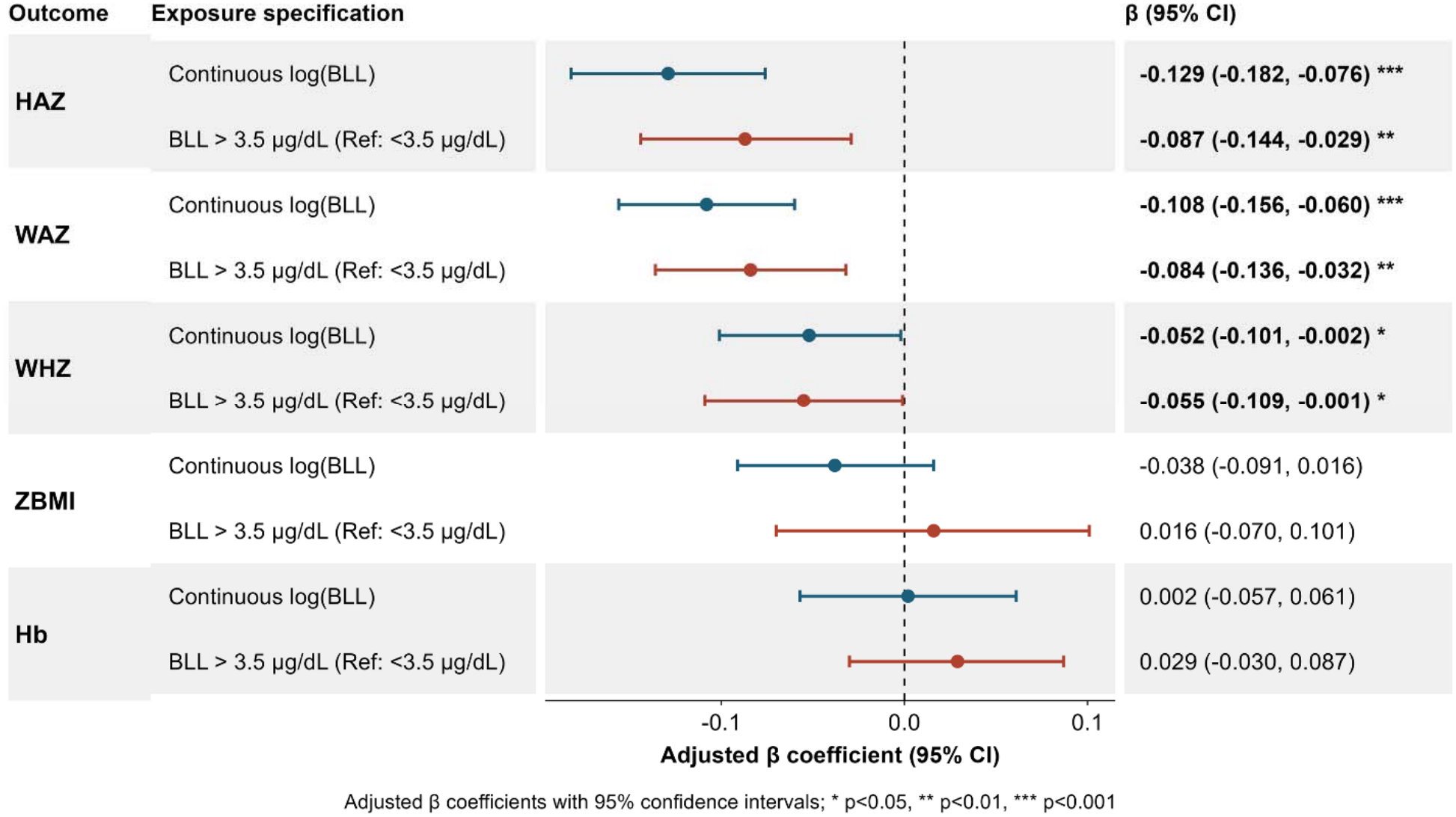
Overall associations of BLL (µg/dL) with child growth and haemoglobin. (**Note:** *P < 0.05, **P<0.01, ***P<0.001. HAZ, height-for-age Z-score; WAZ, weight-for-age Z-score; WHZ, weight-for-height Z-score; ZBMI, body mass index-for-age Z-score; Hb, haemoglobin. Models were adjusted for child sex, child age (months), maternal age (years), maternal education, household wealth index, place of residence, region, water source, sanitation facility, interior wall condition, floor type, household smoking, lead-related activity nearby, and caregiver lead-related occupation).

#### 3.4.2. Sex-specific associations

The inverse associations with linear growth were observed in both boys and girls shown in Figure 3 and Table S3. Among boys, higher log-transformed blood lead concentration was associated with lower HAZ (β = −0.114, 95% CI: −0.188 to −0.040, P < 0.01), WAZ (β = −0.113, 95% CI: −0.179 to −0.046, P < 0.001) and WHZ (β = −0.076, 95% CI: −0.144 to −0.007, P < 0.05). However, no evidence was observed for ZBMI or haemoglobin (P > 0.05). Among girls, higher BLL was associated with lower HAZ (β = −0.150, 95% CI: −0.225 to −0.075, P < 0.001) and WAZ (β = −0.104, 95% CI: −0.171 to −0.036, P = 0.003). No evidence of associations was observed for WHZ, ZBMI or haemoglobin (P > 0.05) among girls. Sensitivity analysis using log_2_-transformed blood lead concentrations yielded similar patterns of associations, supporting the robustness of the primary findings (Table S4).

**Figure 3.**
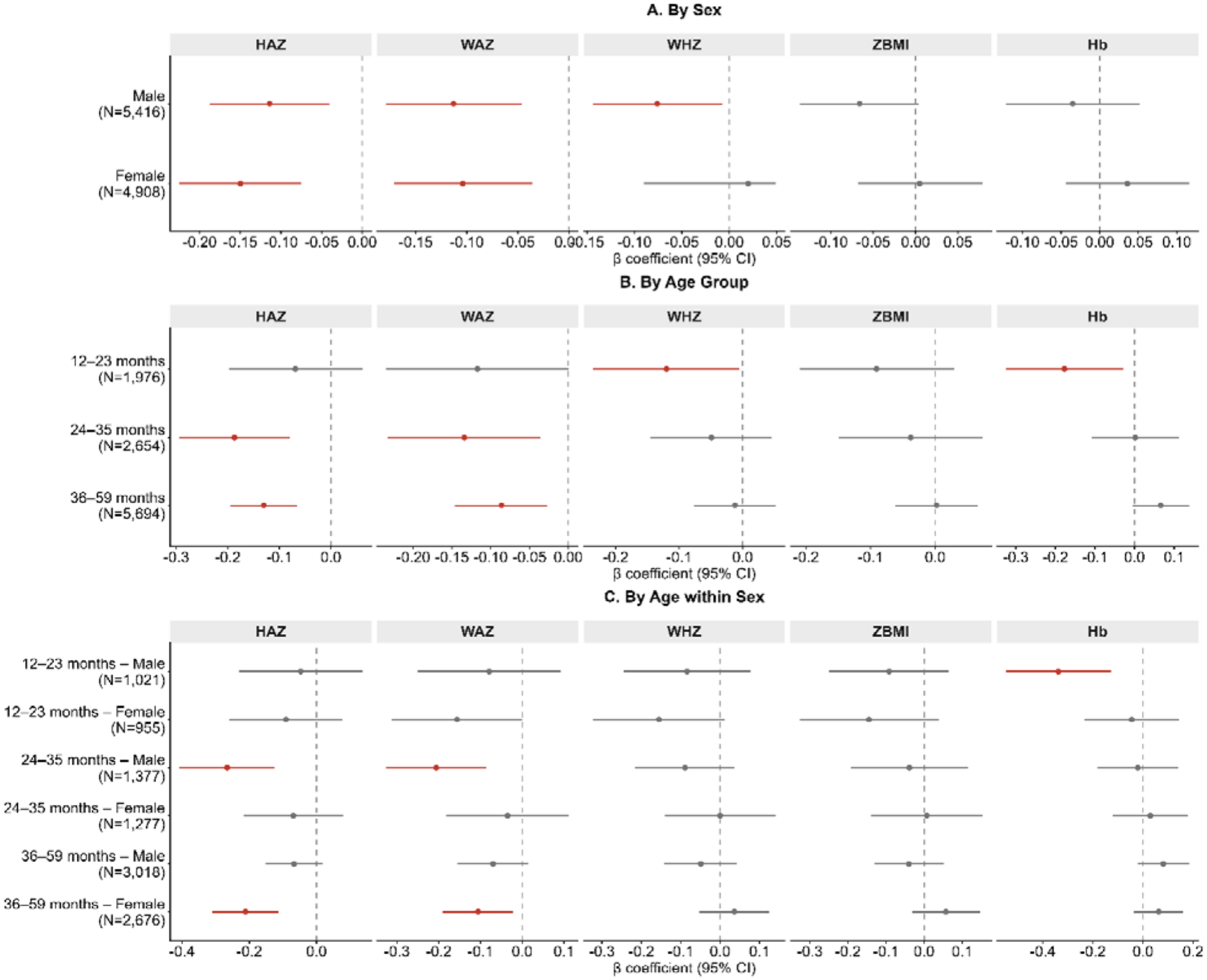
Associations of BLL with child growth and haemoglobin by sex, age group and age within sex. (**Note:** *P < 0.05, **P<0.01, ***P<0.001. HAZ, height-for-age Z-score; WAZ, weight-for-age Z-score; WHZ, weight-for-height Z-score; ZBMI, body mass index-for-age Z-score; Hb, haemoglobin. Models were adjusted for child sex, child age (months), maternal age (years), maternal education, household wealth index, place of residence, region, water source, sanitation facility, interior wall condition, floor type, household smoking, lead-related activity nearby, and caregiver lead-related occupation).

The categorical exposure showed broadly consistent findings. Among boys, BLL of at least 3.5 µg/dL were associated with lower HAZ (β = −0.080, 95% CI: −0.160 to −0.001, P < 0.05), WAZ (β = −0.098, 95% CI: −0.168 to −0.027, P < 0.01), and WHZ (β = −0.086, 95% CI: −0.162 to −0.009, P <0.05) but no associations with ZBMI or haemoglobin (P > 0.05). Among girls, BLL of at least 3.5 µg/dL were associated with lower HAZ (β = −0.095, 95% CI: −0.175 to −0.015, P < 0.05) and WAZ (β = −0.076, 95% CI: −0.150 to −0.002, P < 0.05), but no evidence of associations was observed for WHZ, ZBMI or haemoglobin (P > 0.05) (Figure 3 and Table S3).

#### 3.4.3. Age-specific associations

The associations between BLL and growth measures varied across age groups. Among children aged 12– 23 months, higher log-transformed BLL was associated with lower WHZ (β = −0.120, 95% CI: −0.236 to −0.005, P < 0.05) and lower haemoglobin (β = −0.177, 95% CI: −0.324 to −0.029, P < 0.05), whereas no evidence of associations was observed for HAZ, WAZ or ZBMI (P > 0.05). Among children aged 24–35 months, higher BLL was associated with lower HAZ (β = −0.187, 95% CI −0.294 to −0.080, P < 0.001) and WAZ (β = −0.134, 95% CI: −0.233 to −0.036, P < 0.01). No evidence of associations was observed for WHZ, ZBMI or haemoglobin. Among children aged 36–59 months, higher BLL was associated with lower HAZ (β = −0.130, 95% CI: −0.195 to −0.066, P < 0.001) and WAZ (β = −0.086, 95% CI: −0.146 to −0.027, P < 0.01), whereas no evidence of associations was observed for WHZ, ZBMI or haemoglobin (P > 0.05) (Figure 3 and Table S3). Sensitivity analysis using log_2_-transformed BLL yielded similar patterns of associations, supporting the robustness of the primary findings (Table S4).

The categorical exposure analysis was generally consistent with the continuous exposure analysis, although some associations were attenuated. Among children aged 12–23 months, no statistically significant associations were observed for any outcome. Among those aged 24–35 months, BLL ≥3.5 µg/dL was associated with lower HAZ (β = −0.124, 95% CI: −0.244 to −0.004, P < 0.05), while no evidence of associations was observed for the other outcomes. Among children aged 36–59 months, BLL ≥3.5 µg/dL was associated with lower HAZ (β = −0.079, 95% CI: −0.149 to −0.009, P < 0.05) and WAZ (β = −0.071, 95% CI: −0.142 to −0.001, P < 0.05), with no evidence of associations for WHZ, ZBMI, or haemoglobin (P > 0.05) (Figure 3 and Table S3).

#### 3.4.4. Age within sex associations

Figure 3 and Table S3 shows the age- and sex-specific analyses which indicated a more differentiated pattern of associations across developmental stages and sex. Among boys aged 12–23 months, higher log-transformed BLL was associated with lower haemoglobin (β = −0.338, 95% CI: −0.546 to −0.129, P < 0.01), while no evidence of associations was observed for HAZ, WAZ, WHZ, or ZBMI. The categorical analysis showed a consistent inverse association with haemoglobin (β = −0.233, 95% CI: −0.445 to −0.022, P < 0.05). Among girls aged 12–23 months, no evidence of associations was observed for any of the examined outcomes (P > 0.05).

Among boys aged 24–35 months, the clearest inverse associations with growth were observed for HAZ and WAZ, with higher BLL associated with lower HAZ (β = −0.266, 95% CI: −0.408 to −0.125, P < 0.001) and WAZ (β = −0.206, 95% CI: −0.326 to −0.086, P < 0.001). No evidence of associations was observed for WHZ, ZBMI, or haemoglobin (P > 0.05). The categorical exposure showed a similar pattern for HAZ (β = −0.197, 95% CI: −0.367 to −0.027), but not for WAZ (P > 0.05). Among girls aged 24–35 months, no evidence of associations was observed across the examined outcomes for either exposure specification (P > 0.05).

Among boys aged 36–59 months, no statistically significant associations were observed for any growth or haemoglobin measure. In contrast, among girls aged 36–59 months, higher log-transformed BLL was associated with lower HAZ (β = −0.211, 95% CI: −0.310 to −0.113, P < 0.001) and WAZ (β = −0.106, 95% CI: −0.191 to −0.022, P = 0.014), while no evidence of associations was observed for WHZ, ZBMI, or haemoglobin. Sensitivity analysis using log_2_-transformed BLL yielded similar patterns of associations, supporting the robustness of the primary findings (Table S4). The categorical exposure analysis was consistent for HAZ (β = −0.140, 95% CI: −0.241 to −0.039, P < 0.01) among girls, but the association with WAZ was not statistically significant (P > 0.05) (Figure 3 and Table S3).

### 3.5. Non-linear associations

Restricted cubic spline analyses showed that the associations between BLL and HAZ, WAZ, WHZ, and ZBMI were broadly consistent with linearity in the overall sample, with no evidence of non-linearity for HAZ (P for non-linearity = 0.317), WAZ (P = 0.669), WHZ (P = 0.396), or ZBMI (P = 0.819). In contrast, the association between BLL and haemoglobin showed evidence of non-linearity in the overall sample (P = 0.006) (Figure 4). In sex-specific analyses, no evidence of non-linearity was observed for the growth indicators among either boys or girls, whereas haemoglobin showed evidence of non-linearity among girls (P = 0.047), but not boys (P = 0.152) (Figure S4). Age-specific analyses showed evidence of non-linearity for haemoglobin among children aged 12–23 months (P = 0.005), WHZ among those aged 36–59 months (P = 0.014), and ZBMI among those aged 36–59 months (P = 0.007); the remaining age-specific associations showed no evidence of departure from linearity (Figure S5).

**Figure 4.**
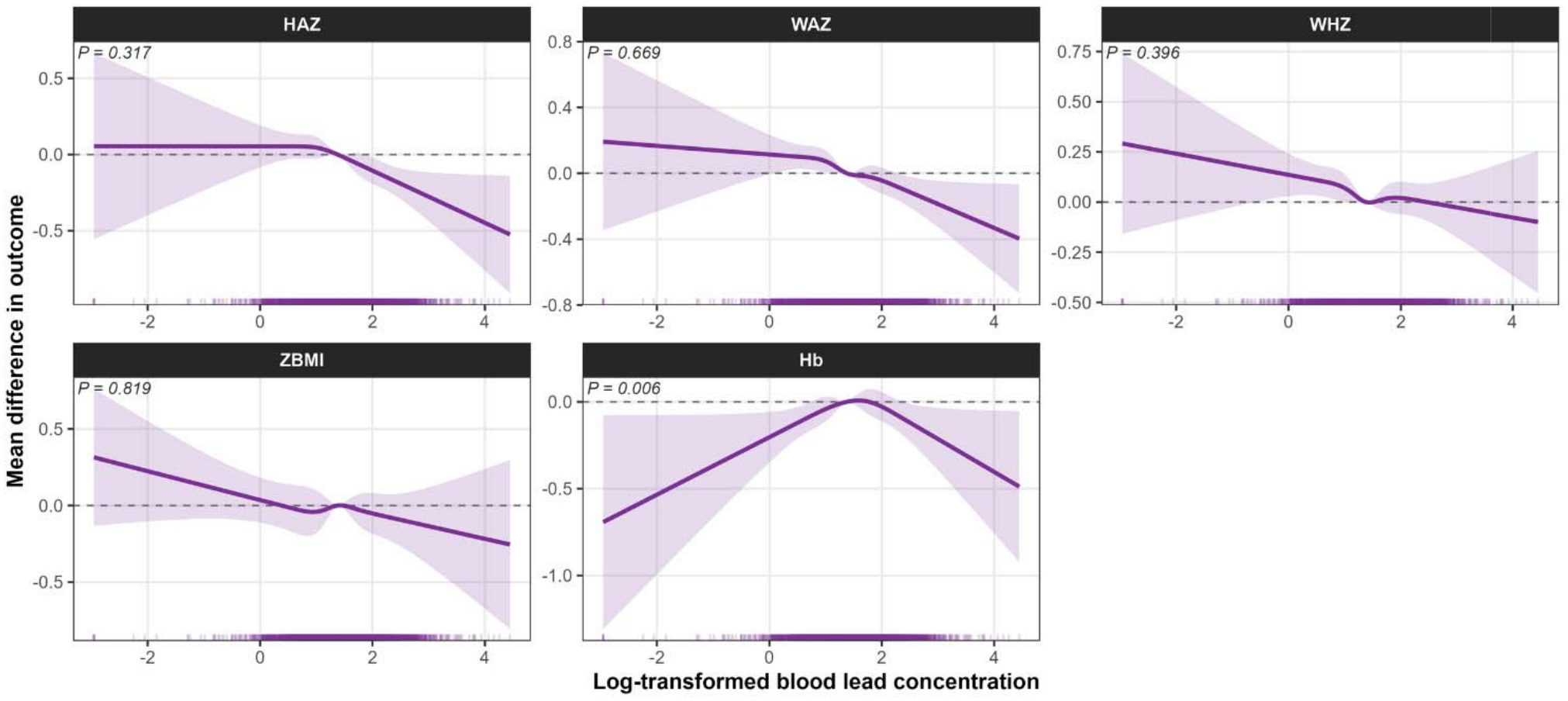
Non-linear associations of BLL with child growth and haemoglobin.

## 4. Discussion

### 4.1. Summary of the main findings

This nationally representative study demonstrated a substantial burden of elevated BLLs among Bangladeshi children aged 12–59 months, with nearly two-thirds (66.26%) exceeding the U.S. CDC BLL reference value of 3.5 µg/dL (CDC, 2021). Against this background, higher BLLs were consistently associated with poorer physical growth, particularly lower HAZ, WAZ, WHZ. The overall pattern wa broadly consistent across boys and girls for HAZ and WAZ, although some outcome-specific difference were apparent. Age-specific analyses further showed that the associations with HAZ and WAZ were more evident among children aged 24–35 and 36–59 months, whereas the association with haemoglobin wa concentrated among children aged 12–23 months, particularly boys. These findings extended previou evidence on childhood lead exposure by examining multiple dimensions of growth within a nationall representative population and by considering whether the observed associations vary acros developmental stages and between boys and girls.

### 4.2. Blood lead levels and child physical growth

Our study found higher BLL to be associated with lower HAZ, suggesting that children with greater lead exposure tended to have poorer linear growth. Consistent with our findings, a study of young Chinese children reported significantly lower HAZ associated with increasing BLLs (Zhou et al., 2020). Similarly, a study of rural Bangladeshi children aged approximately 20–40 months found increased odds of stunting associated with BLL (Gleason et al., 2016). Another study among Bangladeshi slum children reported that elevated BLL was associated with stunting (Raihan et al., 2018). While other studies observed no significant association between BLLs and HAZ (Ahmadi et al., 2022; Taylor et al., 2026).

The association with WAZ showed a similar pattern in our study. Elevated BLL was associated with lower WAZ, indicating that the relationship was not restricted to linear stature but also extended to overall weight relative to age. Previous findings on BLL and childhood weight, however, have been inconsistent. Zhou et al. (2020) for example, found lower WAZ among children with higher BLLs.

Similarly, Raihan et al. (2018) reported an inverse association between higher BLL and WAZ among Bangladeshi slum children. However, other studies including NHANES III and Canadian MIREC cohort, reported no significant association between BLL and body weight (Ashley-Martin et al., 2019; Ballew et al., 1999).

The association with WHZ was also inverse, although weaker and less consistent than those observed for HAZ and WAZ. The weaker association observed for WHZ may therefore reflect the different growth dimension captured by this indicator, which evaluates weight relative to attained height rather than weight or height relative to age. This interpretation is compatible with findings from the Bangladeshi study of young children, which reported an association between elevated BLL and stunting but did not find a significant association with wasting. This study suggested that chronic lead exposure might be more closely related to chronic growth impairment than to the more acute manifestation represented by wasting (Raihan et al., 2018).

These findings might be partly explained by the persistent and heterogeneous nature of lead exposure among Bangladeshi children, whereby repeated exposure may affect growth even at the relatively lower BLLs. In our study, the geometric mean BLL was 4.43 µg/dL, considerably lower than levels reported in earlier Bangladeshi studies (Gleason et al., 2016; Mitra et al., 2012), yet higher BLL was associated with lower HAZ, WAZ and WHZ. Such associations at comparatively lower exposure concentrations suggest that growth-related effects may not be restricted to children with markedly elevated BLLs. Young children have tendency to chew or swallow non-food materials may further increase lead ingestion through contaminated soil, dust, and household materials (De la Burdé and Reames, 1973). Pica, reported in approximately 5.5% of children in rural Bangladesh (Khan et al., 2009), might be major potential exposure pathway. In Bangladesh, other documented sources of childhood lead exposure include industrial contamination, indigenous medicines, paints, and contaminated household materials (Boman et al., 2005; Mitra et al., 2012, 2009).

We found no association between BLL and ZBMI. This finding differs from some previous evidence. Scinicariello et al. (2013), using NHANES data, reported lower BMI associated with higher BLLs, while other studies observed positives (Wang et al., 2015) or null associations between BLL and BMI (Ashley-Martin et al., 2019). The absence of an association with ZBMI in our study should therefore not be interpreted as evidence that BLL is unrelated to growth. Rather, it may reflect the different construct captured by BMI-for-age.

Contrasting with prior studies (Liu et al., 2012; Mitra et al., 2012), our study did not observe association with haemoglobin (Hb). The absence of an overall association in our study may therefore reflect the multifactorial determinants of haemoglobin and the relatively lower exposure distribution. Importantly, however, our spline analysis suggested a non-linear relationship between BLL and Hb, suggesting that the linear estimate may not fully capture the exposure-response pattern. This finding is consistent with the broader toxicological literature showing that dose-response relationships may be non-linear at relatively low exposure levels, although evidence from other outcomes cannot be directly extrapolated to Hb (Lanphear et al., 2005). Therefore, this finding warrants further investigation of non-linearity in the hematological effects of low BLL.

### 4.3. Sex-specific patterns in growth indicators and haemoglobin

Our findings suggest that BLLs and child growth associations were not consistent between boys and girls. Boys had significantly higher BLLs than girls. Despite this exposure differences, elevated BLL was associated with lower HAZ and WAZ in both sexes, whereas the association with WHZ was evident only among boys. These findings are partly consistent with (Zhou et al., 2020), who reported inverse associations of BLL with HAZ and WAZ among young children, with associations more evident among boys. In contrast, the Canadian MIREC cohort reported evidence of sex heterogeneity for the association between BLL and BMI, with higher BLL tending to be associated with lower BMI among girls (Ashley-Martin et al., 2019). The biological basis for the observed sex-specific pattern remains uncertain, but several pathways may plausibly contribute. An earlier review suggested that lead-related toxicity may differ by sex, with some evidence of greater susceptibility among boys (Llop et al., 2013). Vahter et al. (2007) also reported sex-related differences in the disposition and biological effects of toxic metals, and noted that boys may be more susceptible to some developmental effects of BLL. Experimental evidence suggests that male offspring may be more vulnerable to lead-related growth impairment, with developmental lead exposure producing greater suppression of somatic growth in male than female rats (Ronis et al., 1998). Lead may also interfere with endocrine pathways involved in growth, and epidemiological studies have reported sex-specific associations between BLL and thyroid-related measures, including altered thyroid hormone profiles among males (Luo and Hendryx, 2014; Meeker et al., 2009). Although these studies were conducted primarily in adults, these findings suggest that sex-dependent endocrine responses may contribute to differences in growth-related associations.

### 4.4. Differences across developmental stages

Our age-specific findings suggest that the relationship between BLL and child physical growth may vary across developmental stages. The inverse associations with HAZ and WAZ were observed among children aged 24–35 months, and this pattern was mainly observed among boys in this age group. Among girls, the associations were more evident in the 36–59-month group. Previous findings on lead-related growth associations is inconsistent. In the Cincinnati Lead Study, higher BLL during the second and third years of life were associated with lower attained height at 33 months (Shukla et al., 1991). Evidence from Bangladesh is also mixed, with Igra et al. (2021) reporting inverse associations of urinary lead with HAZ and WAZ among boys but not girls at 10 years, whereas an earlier longitudinal study with same population found no significant associations between lead exposure and height or weight through age 5 (Gardner et al., 2013).

For haemoglobin, our study found an inverse association with BLL among 12–23 months aged children and this was more apparent among boys in that age group. Evidence specifically examining age-dependent associations between – and haemoglobin in children is limited. Previous studies have generally examined broader childhood age ranges and have reported inconsistent findings. Mitra et al. (2012) reported an inverse correlation between BLL and haemoglobin among Bangladeshi children younger than 16 years, but did not identify a specific early-childhood age window. Similarly, an Ecuadorian study of children with chronic lead exposure found an overall inverse association between BLL and haemoglobin but no significant age-by-sex differences for haemoglobin (Counter et al., 2000).

The biological basis for the age-specific finding in our study is unclear, but may partly relate to greater gastrointestinal absorption of lead and frequent hand-to-mouth behaviour in young children (CDC, 2025). Children in this age have high physiological iron requirements during the transition to complementary feeding, when iron stores may be vulnerable to depletion (Osendarp et al., 2016). These factors could contribute to the variation observed across age groups, but differences in cumulative exposure, nutritional status, and developmental susceptibility cannot be separated in our cross-sectional study.

### 4.5. Potential mechanisms

Several biological pathways may explain the associations observed between BLL and child growth. Lead may interfere with skeletal growth by affecting bone formation, mineralization, and osteoblast activity. Experimental studies have reported reduced longitudinal bone growth and bone strength following developmental lead exposure (Ronis et al., 2001), while epidemiological evidence has linked higher BLLs with reduced linear skeletal growth (Ignasiak et al., 2006; Li et al., 2022). Lead may also affect the growth hormone (GH)-insulin-like growth factor-1 (IGF-1) axis. In children, higher BLLs have been associated with lower circulating IGF-1 (Fleisch et al., 2013), and experimental studies suggest that lead can interfere with IGF-1 signalling and skeletal growth (Xu et al., 2014). These pathways provide biological support for the inverse associations with HAZ and WAZ observed in our study. Lead can interfere with haem synthesis, while iron deficiency may increase lead absorption, potentially increasing the susceptibility of young children to haematological effects of lead (Capitão et al., 2022; Tempowski, 2021). Lead inhibits key enzymes in the haem biosynthetic pathway, particularly δ-aminolevulinic acid dehydratase and ferrochelatase, thereby impairing haem production (Jangid et al., 2012; Piomelli et al., 1982). Evidence from children has demonstrated inhibition of ALAD with increasing BLLs (Roels et al., 1976), while other evidence supports the relationship between lead exposure and haematological outcomes in children (Rawat et al., 2021). Iron status may further influence this pathway because iron deficiency has been associated with higher BLLs in children (Farkhondeh et al., 2022).

### 4.6 Strengths and limitations

This study has several strengths. It used a large, nationally representative sample with objectively measured BLL, haemoglobin, and anthropometric outcomes. The detailed survey design was incorporated into all regression analyses, and the associations were examined using both continuous and categorical measures of BLL. We also assessed patterns by sex, age, and age-by-sex groups and evaluated potential non-linear associations. However, several limitations should be considered. The cross-sectional design prevents assessment of temporality and causal relationships and does not exclude residual confounding. Important factors such as detailed dietary intake and micronutrient status were not fully available in the dataset. BLL was measured at a single time point, so we could not assess the duration, timing, or cumulative burden of exposure. The anthropometric measures also reflect children’s size at the time of assessment rather than changes in growth over time, while a single haemoglobin measurement cannot establish the underlying cause of low haemoglobin.

## 5. Conclusions

In this nationally representative samples, higher BLLs were associated with lower HAZ, WAZ, and WHZ among Bangladeshi children aged 12–59 months. The sex-specific analyses showed inverse associations of BLL with HAZ and WAZ in both boys and girls, while the association with WHZ was observed only among boys. The age-specific analyses showed associations with HAZ and WAZ among children aged 24–35 and 36–59 months, whereas the association with haemoglobin was observed in 12–23-month group. Further stratification by age within sex showed a more specific pattern, with inverse associations of BLL with HAZ and WAZ among boys aged 24–35 months and girls aged 36–59 months, and with haemoglobin among boys aged 12–23 months. Overall, these findings suggest that the relationship of BLL with child growth and haemoglobin varies across sex and developmental stage, with the most consistent associations observed for linear growth indicators, particularly HAZ and WAZ. Longitudinal studies with repeated measures of BLL, growth, haemoglobin, and nutritional status are needed to determine whether these age- and sex-specific patterns reflect differences in exposure timing, cumulative exposure, or developmental susceptibility.

## Author Contributions

**Muhammad Mainuddin Patwary:** Conceptualization, Data curation, Methodology, Formal analysis, Visualization, Project administration, Supervision, Writing-original draft, Writing-review & editing

**Sadia Ashraf:** Conceptualization, Writing-original draft, Writing-review & editing

**Md. Atikul Islam:** Writing-review & editing

**Matthew HEM Browning:** Writing-review & editing

**Michelle L. Bell:** Writing-review & editing

## Data Availability

This study used publicly available Bangladesh Multiple Indicator Cluster Survey (MICS) 2025 data.

## Consent for publication

All authors consent to publish this article.

## Declaration of Competing Interest

The authors declare that they have no known competing financial interests or personal relationships that could have appeared to influence the work reported in this paper.

## Funding

The study did not receive any funds, grants, or other support.

## Declaration of generative AI and AI-assisted technologies in the manuscript preparation process

During the preparation of this manuscript, the author utilized Claude (Claude Sonnet 5, Anthropic) to assist with language enhancement, clarity, and readability. After employing this tool, the author carefully reviewed and revised the content as necessary and assumed full responsibility for the article’s final version.

## Notes

### Competing Interest Statement

The authors have declared no competing interest.

## References

Abelsohn, A.R., Sanborn, M., 2010. Lead and children: Clinical management for family physicians. Can. Fam. Physician.

Afeiche, M., Peterson, K.E., Sánchez, B.N., Schnaas, L., Cantonwine, D., Ettinger, A.S., Solano-González, M., Hernández-Avila, M., Hu, H., Téllez-Rojo, M.M., 2012. Windows of lead exposure sensitivity, attained height, and body mass index at 48 months. J. Pediatr. 10.1016/j.jpeds.2011.12.022

Ahmadi, S., Botton, J., Zoumenou, R., Ayotte, P., Fievet, N., Massougbodji, A., Alao, M.J., Cot, M., Glorennec, P., Bodeau-Livinec, F., 2022. Lead Exposure in Infancy and Subsequent Growth in Beninese Children. Toxics. 10.3390/toxics10100595

Ashley-Martin, J., Dodds, L., Arbuckle, T.E., Lanphear, B., Muckle, G., Bouchard, M.F., Fisher, M., Asztalos, E., Foster, W., Kuhle, S., 2019. Blood metal levels and early childhood anthropometric measures in a cohort of Canadian children. Environ. Res. 10.1016/j.envres.2019.108736

Ballew, C., Khan, L.K., Kaufmann, R., Mokdad, A., Miller, D.T., Gunter, E.W., 1999. Blood lead concentration and children’s anthropometric dimensions in the Third National Health and Nutrition Examination Survey (NHANES III), 1988-1994. J. Pediatr. 10.1016/S0022-3476(99)70250-7

BBS, B.B. of S., UNICEF, 2026. Bangladesh Multiple Indicator Cluster Survey 2025 | UNICEF Bangladesh [WWW Document]. URL https://www.unicef.org/bangladesh/en/media/13106/file/Bangladesh 2025 MICS Survey Findings Report.pdf.pdf (accessed 9.28.26).

Boman, J., Gatari, M.J., Wagner, A., Hossain, M.I., 2005. Elemental characterization of aerosols in urban and rural locations in Bangladesh. X-Ray Spectrom. 10.1002/xrs.864

Capitão, C., Martins, R., Santos, O., Bicho, M., Szigeti, T., Katsonouri, A., Bocca, B., Ruggieri, F., Wasowicz, W., Tolonen, H., Virgolino, A., 2022. Exposure to heavy metals and red blood cell parameters in children: A systematic review of observational studies. Front. Pediatr. 10.3389/fped.2022.921239

CDC, 2025. Chapter 3. Toxicokinetics, Susceptible Populations,Biomarkers, Cemical Interactions [WWW Document]. Agency Toxic Subst. Dis. Regist.

CDC, 2021. CDC Updates Blood Lead Reference Value | Childhood Lead Poisoning Prevention | CDC [WWW Document]. URL https://www.cdc.gov/lead-prevention/php/news-features/updates-blood-lead-reference-value.html (accessed 9.28.26).

Counter, S.A., Buchanan, L.H., Ortega, F., Rifai, N., 2000. Blood lead and hemoglobin levels in Andean children with chronic lead intoxication. Neurotoxicology.

De la Burdé, B., Reames, B., 1973. Prevention of pica, the major cause of lead poisoning in children. Am. J. Public Health. 10.2105/AJPH.63.8.737

Ericson, B., Hu, H., Nash, E., Ferraro, G., Sinitsky, J., Taylor, M.P., 2021. Blood lead levels in low-income and middle-income countries: a systematic review. Lancet Planet. Heal. 10.1016/S2542-5196(20)30278-3

Farkhondeh, T., Mansouri, B., Binkowski, L.J., Błaszczyk, M., Pirsaheb, M., Azadi, N.A., Słoboda, M., Amirabadizadeh, A., Javadmoosavi, S.Y., 2022. Blood lead concentrations in children with iron deficiency anemia: a systematic review and meta-analysis. Environ. Sci. Pollut. Res. 10.1007/s11356-021-17301-z

Fleisch, A.F., Burns, J.S., Williams, P.L., Lee, M.M., Sergeyev, O., Korrick, S.A., Hauser, R., 2013. Blood lead levels and serum insulin-like growth factor 1 concentrations in peripubertal boys. Environ. Health Perspect. 10.1289/ehp.1206105

Gardner, R.M., Kippler, M., Tofail, F., Bottai, M., Hamadani, J., Grandér, M., Nermell, B., Palm, B., Rasmussen, K.M., Vahter, M., 2013. Environmental exposure to metals and children’s growth to age 5 years: A prospective cohort study. Am. J. Epidemiol. 10.1093/aje/kws437

Gleason, K.M., Valeri, L., Shankar, A.H., Hasan, M.O.S.I., Quamruzzaman, Q., Rodrigues, E.G., Christiani, D.C., Wright, R.O., Bellinger, D.C., Mazumdar, M., 2016. Stunting is associated with blood lead concentration among Bangladeshi children aged 2-3 years. Environ. Heal. A Glob. Access Sci. Source. 10.1186/s12940-016-0190-4

Hauptman, M., Bruccoleri, R., Woolf, A.D., 2017. An Update on Childhood Lead Poisoning. Clin. Pediatr. Emerg. Med. 10.1016/j.cpem.2017.07.010

Ignasiak, Z., Sławińska, T., Rozek, K., Little, B.B., Malina, R.M., 2006. Lead and growth status of schoolchildren living in the copper basin of south-western Poland: Differential effects on bone growth. Ann. Hum. Biol. 10.1080/03014460600730752

Irawati, Y., Kusnoputranto, H., Achmadi, U.F., Safrudin, A., Sitorus, A., Risandi, R., Wangsamuda, S., Asih, P.B.S., Syafruddin, D., 2022. Blood lead levels and lead toxicity in children aged 1-5 years of Cinangka Village, Bogor Regency. PLoS One. 10.1371/journal.pone.0264209

Jangid, A.P., John, P.J., Yadav, D., Mishra, S., Sharma, P., 2012. Impact of chronic lead exposure on selected biological markers. Indian J. Clin. Biochem. 10.1007/s12291-011-0163-x

Khan, N.Z., Ferdous, S., Islam, R., Sultana, A., Durkin, M., Mcconachie, H., 2009. Behaviour problems in young children in rural Bangladesh. J. Trop. Pediatr. 10.1093/tropej/fmn108

Lanphear, B.P., Hornung, R., Khoury, J., Yolton, K., Baghurst, P., Bellinger, D.C., Canfield, R.L., Dietrich, K.N., Bornschein, R., Greene, T., Rothenberg, S.J., Needleman, H.L., Schnaas, L., Wasserman, G., Graziano, J., Roberts, R., 2005. Low-level environmental lead exposure and children’s intellectual function: An international pooled analysis. Environ. Health Perspect. 10.1289/ehp.7688

Larsen, B., Sánchez-Triana, E., 2023. Global health burden and cost of lead exposure in children and adults: a health impact and economic modelling analysis. Lancet Planet. Heal. 10.1016/S2542-5196(23)00166-3

Li, T., Xie, Y., Wang, L., Huang, G., Cheng, Y., Hou, D., Liu, W., Zhang, T., Liu, J., 2022. The Association between Lead Exposure and Bone Mineral Density in Childhood and Adolescence: Results from NHANES 1999–2006 and 2011–2018. Nutrients. 10.3390/nu14071523

Liu, J., McCauley, L., Yan, C., Shen, X., Pinto-Martin, J.A., 2012. Low blood lead levels and hemoglobin concentrations in preschool children in China. Toxicol. Environ. Chem. 10.1080/02772248.2011.628001

Llop, S., Lopez-Espinosa, M.J., Rebagliato, M., Ballester, F., 2013. Gender differences in the neurotoxicity of metals in children. Toxicology. 10.1016/j.tox.2013.04.015

Luo, J., Hendryx, M., 2014. Relationship between blood cadmium, lead, and serum thyroid measures in US adults - The National Health and Nutrition Examination Survey (NHANES) 2007-2010. Int. J. Environ. Health Res. 10.1080/09603123.2013.800962

Malin Igra, A., Warnqvist, A., Rahman, S.M., Ekström, E.C., Rahman, A., Vahter, M., Kippler, M., 2021. Environmental metal exposure and growth to 10 years of age in a longitudinal mother–child cohort in rural Bangladesh. Environ. Int. 10.1016/j.envint.2021.106738

Meeker, J.D., Rossano, M.G., Protas, B., Diamond, M.P., Puscheck, E., Daly, D., Paneth, N., Wirth, J.J., 2009. Multiple metals predict prolactin and thyrotropin (TSH) levels in men. Environ. Res. 10.1016/j.envres.2009.06.004

Mitra, A.K., Ahua, E., Sana, P.K., 2012. Prevalence of and risk factors for lead poisoning in young children in Bangladesh. J. Heal. Popul. Nutr. 10.3329/jhpn.v30i4.13292

Mitra, A.K., Haque, A., Islam, M., Bashar, S.A.M.K., 2009. Lead poisoning: An alarming public health problem in bangladesh. Int. J. Environ. Res. Public Health. 10.3390/ijerph6010084

Osendarp, S.J.M., Broersen, B., Van Liere, M.J., De-Regil, L.M., Bahirathan, L., Klassen, E., Neufeld, L.M., 2016. Complementary Feeding Diets Made of Local Foods Can Be Optimized, but Additional Interventions Will Be Needed to Meet Iron and Zinc Requirements in 6-to 23-Month-Old Children in Low-and Middle-Income Countries. Food Nutr. Bull. 10.1177/0379572116655239

Piomelli, S., Seaman, C., Zullow, D., Curran, A., Davidow, B., 1982. Threshold for lead damage to heme synthesis in urban children. Proc. Natl. Acad. Sci. U. S. A. 10.1073/pnas.79.10.3335

Raihan, M.J., Briskin, E., Mahfuz, M., Islam, M.M., Mondal, D., Hossain, M.I., Ahmed, A.M.S., Haque, R., Ahmed, T., 2018. Examining the relationship between blood lead level and stunting, wasting and underweight-A cross-sectional study of children under 2 years-of-age in a Bangladeshi slum. PLoS One. 10.1371/journal.pone.0197856

Rawat, P.S., Singh, S., Zahid, M., Mehrotra, S., 2021. An integrated assessment of lead exposure in children: Correlation with biochemical and haematological indices. J. Trace Elem. Med. Biol. 10.1016/j.jtemb.2021.126835

Roels, H., Buchet, J.P., Lauwerys, R., 1976. Impact of air pollution by lead on the heme biosynthetic pathway in school-age children. Arch. Environ. Health. 10.1080/00039896.1976.10667241

Ronis, M.J.J., Aronson, J., Gao, G.G., Hogue, W., Skinner, R.A., Badger, T.M., Lumpkin, C.K., 2001. Skeletal effects of developmental lead exposure in rats. Toxicol. Sci. 10.1093/toxsci/62.2.321

Ronis, M.J.J., Badger, T., Shema, S.J., Roberson, P.K., Templer, L., Ringer, D., Thomas, P.E., 1998.] Endocrine mechanisms underlying the growth effects of developmental lead exposure in the rat. J. Toxicol. Environ. Heal. - Part A. 10.1080/009841098158944

Roy, R., Das, R., 2026. Blood lead levels (BLLs), exposure pathways and health impacts in South Asia: a comprehensive review. Crit. Rev. Toxicol. 10.1080/10408444.2026.2692126

Rutstein, S.O.G., Rojas, M.C.S., 2006. GUIDE TO DHS STATISTICS.

Shukla, R., Dietrich, K.N., Bornschein, R.L., Berger, O., Hammond, P.B., 1991. Lead exposure and growth in the early preschool child: A follow-up report from the Cincinnati lead study. Pediatrics. 10.1542/peds.88.5.886

Taylor, C.M., Golding, J., Kordas, K., 2026. Prenatal lead exposure: associations with growth and anthropometry in early childhood in a UK observational birth cohort study. Wellcome Open Res. 10.12688/wellcomeopenres.16338.3

Tempowski, J., 2021. WHO guideline for clinical management of exposure to lead, World Health Organization.

UNICEF, Pure Earth, 2020. The toxic truth | UNICEF [WWW Document]. URL https://www.unicef.org/reports/toxic-truth-childrens-exposure-to-lead-pollution-2020 (accessed 9.30.26).

Wang, N., Chen, C., Nie, X., Han, B., Li, Q., Chen, Yi, Zhu, C., Chen, Yingchao, Xia, F., Cang, Z., Lu, M., Meng, Y., Zhai, H., Lin, D., Cui, S., Jensen, M.D., Lu, Y., 2015. Blood lead level and its association with body mass index and obesity in China - Results from SPECT-China study. Sci. Rep. 10.1038/srep18299

WHO, W.H.O., 2026. Lead poisoning [WWW Document]. URL https://www.who.int/news-room/fact-sheets/detail/lead-poisoning-and-health (accessed 9.30.26).

WHO, W.H.O., 2025. Progress on household drinking water, sanitation and hygiene 2000-2024: special focus on inequalities. World Heal. Organ.

WHO, W.H.O., 2011. Haemoglobin concentrations for the diagnosis of anaemia and assessment of severity [WWW Document]. URL https://www.who.int/publications/i/item/WHO-NMH-NHD-MNM-11.1 (accessed 9.29.26).

Xu, Y., Liu, M.C., Wang, P., Xu, B., Liu, X.Q., Zhang, Z.P., Ren, L.F., Qin, Q., Ma, Y.Y., Luo, W.J., Hao, X.K., 2014. Correlation between serum IGF-1 and blood lead level in short stature children and adolescent with growth hormone deficiency. Int. J. Clin. Exp. Med.

Zhou, C.C., He, Y.Q., Gao, Z.Y., Wu, M.Q., Yan, C.H., 2020. Sex differences in the effects of lead exposure on growth and development in young children. Chemosphere. 10.1016/j.chemosphere.2020.126294

