## Supplementary files for "Sex- and Age-Specific Associations of Blood Lead Levels with Child Growth and Haemoglobin in Bangladesh: A Nationally Representative Study"

***Corresponding author:**

Muhammad Mainuddin Patwary

**Table S1.** Variance inflation factors (VIFs) for predictors included in the multivariable regression model.

| **Predictor** | **GVIF** | **df** | **Adjusted GVIF** |
| --- | --- | --- | --- |
| Blood lead level (µg/dL) | 1.277 | 1 | 1.13 |
| Child sex | 1.01 | 1 | 1.005 |
| Child age (months) | 1.047 | 1 | 1.023 |
| Maternal age | 1.143 | 1 | 1.069 |
| Maternal education | 1.447 | 5 | 1.038 |
| Household wealth index | 3.271 | 4 | 1.16 |
| Place of residence | 1.2 | 1 | 1.096 |
| Region | 1.688 | 7 | 1.038 |
| Interior wall condition | 1.514 | 2 | 1.109 |
| Floor type | 2.382 | 2 | 1.242 |
| Drinking water source | 1.037 | 2 | 1.009 |
| Sanitation facility | 1.054 | 2 | 1.013 |
| Household smoking | 1.055 | 1 | 1.027 |
| Any lead-related activity nearby | 1.157 | 1 | 1.076 |
| Caregiver engaged in lead-related work | 1.12 | 1 | 1.058 |

GVIF = generalized variance inflation factor; df = degrees of freedom

**Table S2**. Distribution of children by BLL (µg/dL) category, sex, and age group.

| **Distribution** | **N** | **<3.5 µg/dL, n (%)** | **≥3.5 µg/dL, n (%)** | **P-value** |
| --- | --- | --- | --- | --- |
| **Overall** |  |  |  |  |
| **All children** | 10,324 | 4,107 (33.74) | 6,217 (66.26) | — |
| Boys | 5,416 | 2,043 (31.52) | 3,373 (68.48) | <0.001*** |
| Girls | 4,908 | 2,064 (36.20) | 2,844 (63.80) |  |
| **Age group** |  |  |  |  |
| 12–23 months | 1,976 | 685 (29.32) | 1,291 (70.68) | <0.001*** |
| 24–35 months | 2,654 | 1,032 (33.91) | 1,622 (66.09) |  |
| 36–59 months | 5,694 | 2,390 (35.20) | 3,304 (64.80) |  |
| **Age within sex** |  |  |  |  |
| **Boys** |  |  |  |  |
| 12–23 months | 1,021 | 360 (29.20) | 661 (70.80) | 0.269 |
| 24–35 months | 1,377 | 515 (32.79) | 862 (67.21) |  |
| 36–59 months | 3,018 | 1,168 (31.74) | 1,850 (68.26) |  |
| **Girls** |  |  |  |  |
| 12–23 months | 955 | 325 (29.45) | 630 (70.55) | <0.001*** |
| 24–35 months | 1,277 | 517 (35.06) | 760 (64.94) |  |
| 36–59 months | 2,676 | 1,222 (39.19) | 1,454 (60.81) |  |

**Table S3.** Association between BLL and child growth indicators and haemoglobin levels.

|  | **N** | **Exposure** | **HAZ** | **WAZ** | **WHZ** | **ZBMI** | **Hb** |
| --- | --- | --- | --- | --- | --- | --- | --- |
| **Overall** | 10,324 | log-BLL | −0.129 (−0.182, −0.076)*** | −0.108 (−0.156, −0.060)*** | −0.052 (−0.101, −0.002)* | −0.038 (−0.091, 0.016) | 0.002 (−0.057, 0.061) |
|  |  | BLL ≥3.5 µg/dL  (Ref: <3.5 µg/dL) | −0.087 (−0.144, −0.029)** | −0.084 (−0.136, −0.032)** | −0.055 (−0.109, −0.001)* | 0.016 (−0.070, 0.101) | 0.029 (−0.030, 0.087) |
| **By Sex** | | | | | | | |
| Boys | 5,416 | log-BLL | −0.114 (−0.188, −0.040)** | −0.113 (−0.179, −0.046)*** | −0.076 (−0.144, −0.007)* | −0.066 (−0.137, 0.004) | −0.035 (−0.121, 0.052) |
|  |  | BLL ≥3.5 µg/dL  (Ref: <3.5 µg/dL) | −0.080 (−0.160, −0.001)* | −0.098 (−0.168, −0.027)** | −0.086 (−0.162, −0.009)* | 0.013 (−0.130, 0.155) | −0.004 (−0.089, 0.082) |
| Girls | 4,908 | log-BLL | −0.150 (−0.225, −0.075)*** | −0.104 (−0.171, −0.036)** | 0.020 (−0.090, 0.049) | 0.005 (−0.068, 0.079) | 0.036 (−0.044, 0.116) |
|  |  | BLL ≥3.5 µg/dL  (Ref: <3.5 µg/dL) | −0.095 (−0.175, −0.015)* | −0.076 (−0.150, −0.002)* | −0.027 (−0.102, 0.047) | 0.025 (−0.084, 0.134) | 0.058 (−0.024, 0.140) |
| **By Age group** | | | | | | | |
| 12–23 months | 1,976 | log-BLL | −0.069 (−0.198, 0.061) | −0.117 (−0.235, 0.001) | −0.120 (−0.236, −0.005)* | −0.091 (−0.210, 0.029) | −0.177 (−0.324, −0.029)* |
|  |  | BLL ≥3.5 µg/dL  (Ref: <3.5 µg/dL) | −0.078 (−0.235, 0.079) | −0.102 (−0.232, 0.029) | −0.092 (−0.224, 0.039) | 0.164 (−0.185, 0.513) | −0.117 (−0.262, 0.029) |
| 24–35 months | 2,654 | log-BLL | −0.187 (−0.294, −0.080)*** | −0.134 (−0.233, −0.036)** | −0.049 (−0.145, 0.046) | −0.038 (−0.149, 0.073) | 0.002 (−0.108, 0.113) |
|  |  | BLL ≥3.5 µg/dL  (Ref: <3.5 µg/dL) | −0.124 (−0.244, −0.004)* | −0.098 (−0.197, 0.001) | −0.049 (−0.149, 0.051) | 0.047 (−0.149, 0.243) | 0.043 (−0.068, 0.155) |
| 36–59 months | 5,694 | log-BLL | −0.130 (−0.195, −0.066)*** | −0.086 (−0.146, −0.027)** | −0.012 (−0.077, 0.052) | 0.002 (−0.062, 0.066) | 0.066 (−0.005, 0.138) |
|  |  | BLL ≥3.5 µg/dL  (Ref: <3.5 µg/dL) | −0.079 (−0.149, −0.009)* | −0.071 (−0.142, −0.001)* | −0.037 (−0.108, 0.033) | −0.029 (−0.098, 0.040) | 0.061 (−0.009, 0.131) |
| **Age within sex** | | | | | | | |
| Boys (12–23 months) | 1,021 | log-BLL | −0.047 (−0.230, 0.136) | −0.079 (−0.250, 0.092) | −0.084 (−0.245, 0.077) | −0.092 (−0.249, 0.065) | −0.338 (−0.546, −0.129)** |
|  |  | BLL ≥3.5 µg/dL  (Ref: <3.5 µg/dL) | −0.073 (−0.290, 0.144) | −0.098 (−0.277, 0.081) | −0.098 (−0.281, 0.085) | 0.173 (−0.327, 0.674) | −0.233 (−0.445, −0.022)* |
| Boys (24–35 months) | 1,377 | log-BLL | −0.266 (−0.408, −0.125)*** | −0.206 (−0.326, −0.086)*** | −0.089 (−0.216, 0.037) | −0.039 (−0.193, 0.116) | −0.021 (−0.181, 0.140) |
|  |  | BLL ≥3.5 µg/dL  (Ref: <3.5 µg/dL) | −0.197 (−0.367, −0.027)* | −0.111 (−0.248, 0.026) | −0.013 (−0.157, 0.132) | 0.207 (−0.217, 0.631) | 0.000 (−0.171, 0.172) |
| Boys (36–59 months) | 3,018 | log-BLL | −0.067 (−0.152, 0.018) | −0.070 (−0.155, 0.015) | −0.049 (−0.142, 0.043) | −0.040 (−0.130, 0.051) | 0.081 (−0.022, 0.184) |
|  |  | BLL ≥3.5 µg/dL  (Ref: <3.5 µg/dL) | −0.020 (−0.116, 0.075) | −0.069 (−0.163, 0.025) | −0.088 (−0.188, 0.013) | −0.088 (−0.187, 0.011) | 0.095 (−0.004, 0.195) |
| Girls (12–23 months) | 955 | log-BLL | −0.091 (−0.259, 0.077) | −0.156 (−0.312, 0.000) | −0.155 (−0.323, 0.012) | −0.145 (−0.327, 0.038) | −0.045 (−0.233, 0.143) |
|  |  | BLL ≥3.5 µg/dL  (Ref: <3.5 µg/dL) | −0.077 (−0.301, 0.146) | −0.092 (−0.281, 0.097) | −0.071 (−0.259, 0.117) | 0.176 (−0.305, 0.658) | −0.037 (−0.237, 0.162) |
| Girls (24–35 months) | 1,277 | log-BLL | −0.069 (−0.217, 0.079) | −0.035 (−0.182, 0.112) | 0.000 (−0.141, 0.141) | 0.007 (−0.139, 0.153) | 0.029 (−0.121, 0.178) |
|  |  | BLL ≥3.5 µg/dL  (Ref: <3.5 µg/dL) | −0.003 (−0.163, 0.157) | −0.044 (−0.179, 0.091) | −0.065 (−0.199, 0.069) | −0.069 (−0.212, 0.073) | 0.111 (−0.049, 0.271) |
| Girls (36–59 months) | 2,676 | log-BLL | −0.211 (−0.310, −0.113)*** | −0.106 (−0.191, −0.022)* | 0.036 (−0.053, 0.125) | 0.057 (−0.032, 0.147) | 0.062 (−0.036, 0.161) |
|  |  | BLL ≥3.5 µg/dL  (Ref: <3.5 µg/dL) | −0.140 (−0.241, −0.039)** | −0.069 (−0.166, 0.028) | 0.025 (−0.070, 0.120) | 0.042 (−0.052, 0.136) | 0.038 (−0.058, 0.135) |

**Note:** *P < 0.05, **P<0.01, ***P<0.001.

BLL, Blood Lead Level; HAZ, height-for-age Z-score; WAZ, weight-for-age Z-score; WHZ, weight-for-height Z-score; ZBMI, body mass index-for-age Z-score; Hb, haemoglobin.

Models were adjusted for child sex, child age (months), maternal age (years), maternal education, household wealth index, place of residence, region, water source, sanitation facility, interior wall condition, floor type, household smoking, lead-related activity nearby, and caregiver lead-related occupation.

**Table S4.** Sensitivity analysis on the association between BLL using log2 and child growth indicators and haemoglobin levels.

|  | **N** | **Exposure** | **HAZ** | **WAZ** | **WHZ** | **ZBMI** | **Hb** |
| --- | --- | --- | --- | --- | --- | --- | --- |
| All children | 10,324 | log2 BLL | −0.089 (−0.126, −0.052)*** | −0.075 (−0.108, −0.042)*** | −0.036 (−0.070, −0.002)* | −0.026 (−0.063, 0.011) | 0.001 (−0.040, 0.042) |
| **By sex** | | | | | | | |
| Boys | 5,416 | log2 BLL | −0.079 (−0.130, −0.028)** | −0.078 (−0.124, −0.032)*** | −0.053 (−0.100, −0.005)* | −0.046 (−0.095, 0.003) | −0.024 (−0.084, 0.036) |
| Girls | 4,908 | log2 BLL | −0.104 (−0.156, −0.052)** | −0.072 (−0.118, −0.025)** | −0.014 (−0.062, 0.034) | 0.004 (−0.047, 0.055) | 0.025 (−0.030, 0.081) |
| **By Age group** | | | | | | | |
| 12-23m | 1,976 | log2 BLL | −0.048 (−0.137, 0.042) | −0.081 (−0.163, 0.001) | −0.083 (−0.164, −0.003)* | −0.063 (−0.146, 0.020) | −0.122 (−0.225, −0.020)* |
| 24-35m | 2,654 | log2 BLL | −0.130 (−0.203, −0.056)*** | −0.093 (−0.162, −0.025)** | −0.034 (−0.101, 0.032) | −0.026 (−0.104, 0.051) | 0.002 (−0.075, 0.078) |
| 36-59m | 5,694 | log2 BLL | −0.090 (−0.135, −0.046)*** | −0.060 (−0.101, −0.018)** | −0.009 (−0.053, 0.036) | 0.002 (−0.043, 0.046) | 0.046 (−0.004, 0.096) |
| **By Age within sex** | | | | | | | |
| Boys (12–23 months) | 1,021 | log2 BLL | −0.033 (−0.160, 0.094) | −0.055 (−0.173, 0.064) | −0.058 (−0.170, 0.054) | −0.064 (−0.172, 0.045) | −0.234 (−0.379, −0.089)** |
| Boys (24–35 months) | 1,377 | log2 BLL | −0.185 (−0.283, −0.087)*** | −0.143 (−0.226, −0.060)*** | −0.062 (−0.150, 0.026) | −0.027 (−0.134, 0.080) | −0.014 (−0.126, 0.097) |
| Boys (36–59 months) | 3,018 | log2 BLL | −0.046 (−0.106, 0.013) | −0.048 (−0.107, 0.011) | −0.034 (−0.098, 0.030) | −0.028 (−0.090, 0.035) | 0.056 (−0.015, 0.128) |
| Girls (12–23 months) | 955 | log2 BLL | −0.063 (−0.179, 0.053) | −0.108 (−0.216, 0.000) | −0.108 (−0.224, 0.009) | −0.100 (−0.227, 0.026) | −0.031 (−0.162, 0.099) |
| Girls (24–35 months) | 1,277 | log2 BLL | −0.048 (−0.150, 0.055) | −0.024 (−0.126, 0.078) | 0.000 (−0.098, 0.098) | 0.005 (−0.096, 0.106) | 0.020 (−0.084, 0.123) |
| Girls (36–59 months) | 2,676 | log2 BLL | −0.147 (−0.215, −0.078)*** | −0.074 (−0.132, −0.015)* | 0.025 (−0.037, 0.087) | 0.040 (−0.022, 0.102) | 0.043 (−0.025, 0.112) |

**Note:** *P < 0.05, **P<0.01, ***P<0.001.

BLL, Blood Lead Level; HAZ, height-for-age Z-score; WAZ, weight-for-age Z-score; WHZ, weight-for-height Z-score; ZBMI, body mass index-for-age Z-score; Hb, haemoglobin.

Models were adjusted for child sex, child age (months), maternal age (years), maternal education, household wealth index, place of residence, region, water source, sanitation facility, interior wall condition, floor type, household smoking, lead-related activity nearby, and caregiver lead-related occupation.

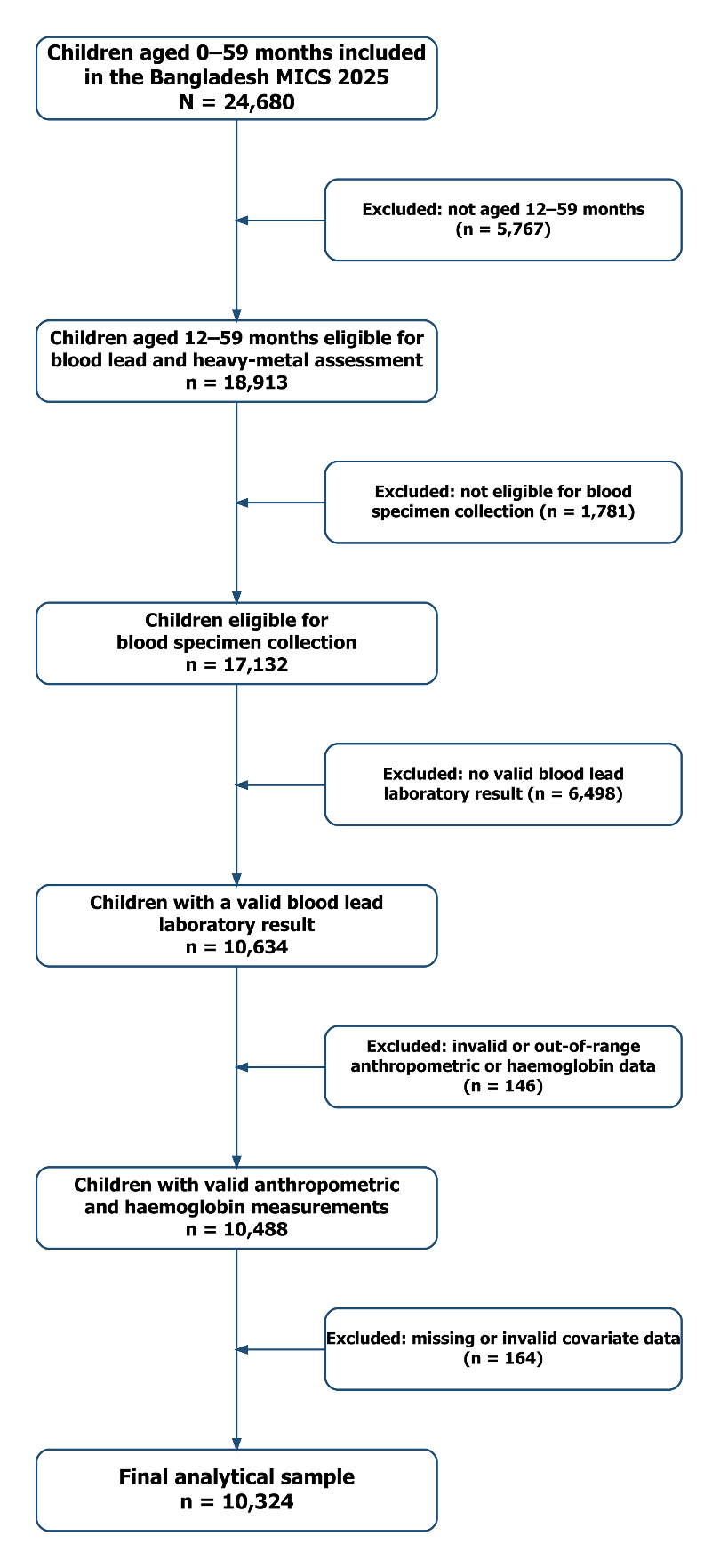

**Figure S1**. Participants flow chart.

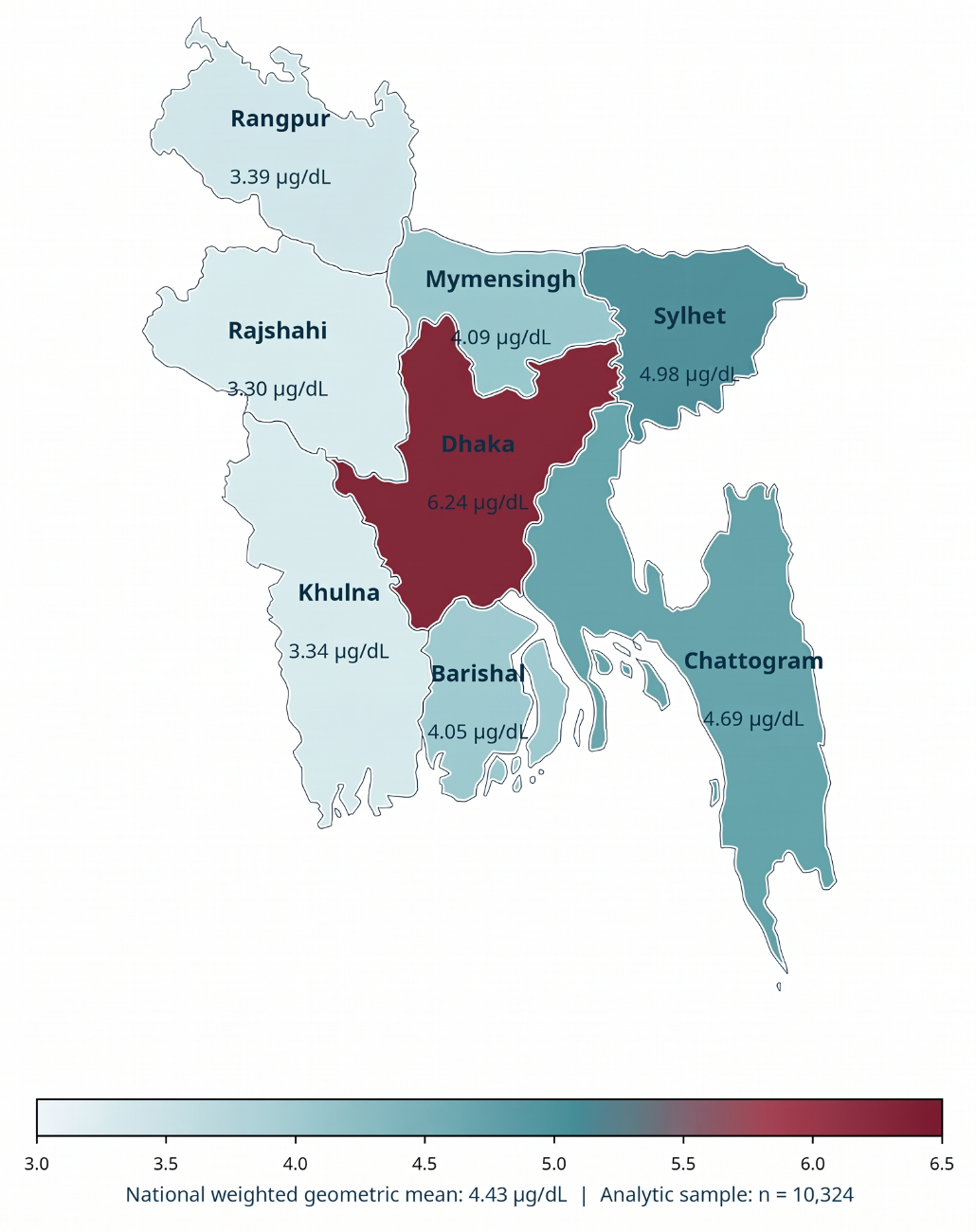

**Figure S2.** Spatial distributions of BLL (µg/dL) among Bangladeshi children aged 12−59 months.

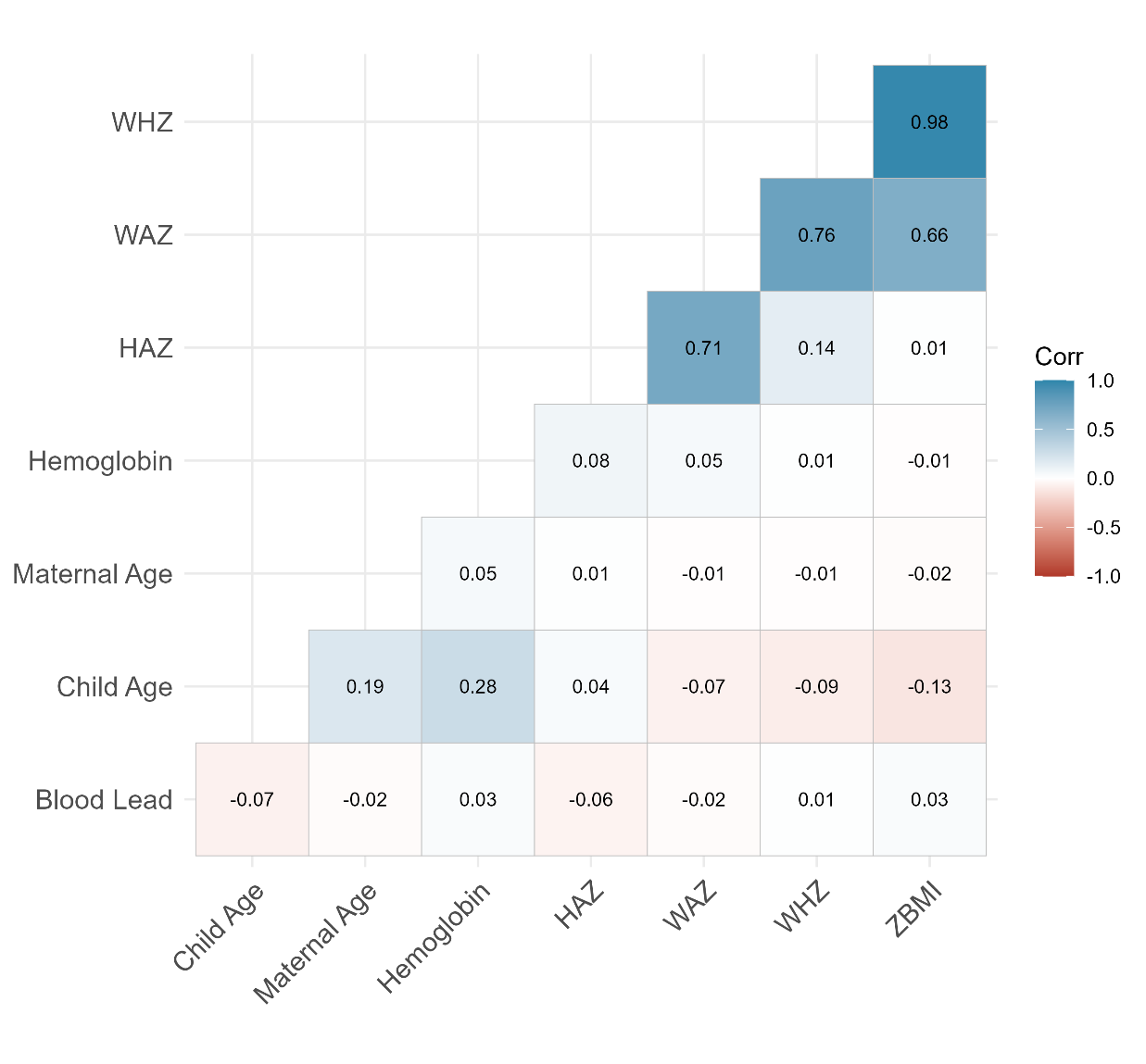

**Figure S3.** Spearman correlations of continuous predictors, exposure and outcomes.

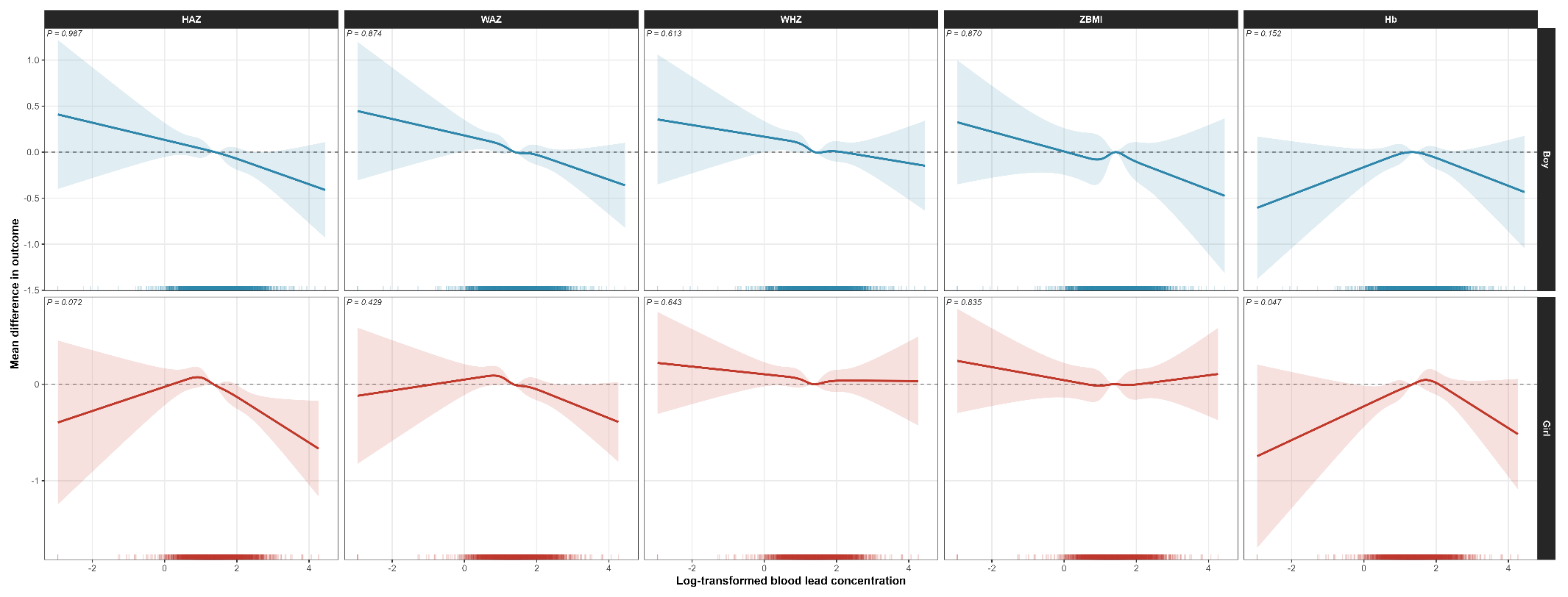

**Figure S4.** Non-linear associations of BLL (µg/dL) with child growth and haemoglobin by sex.

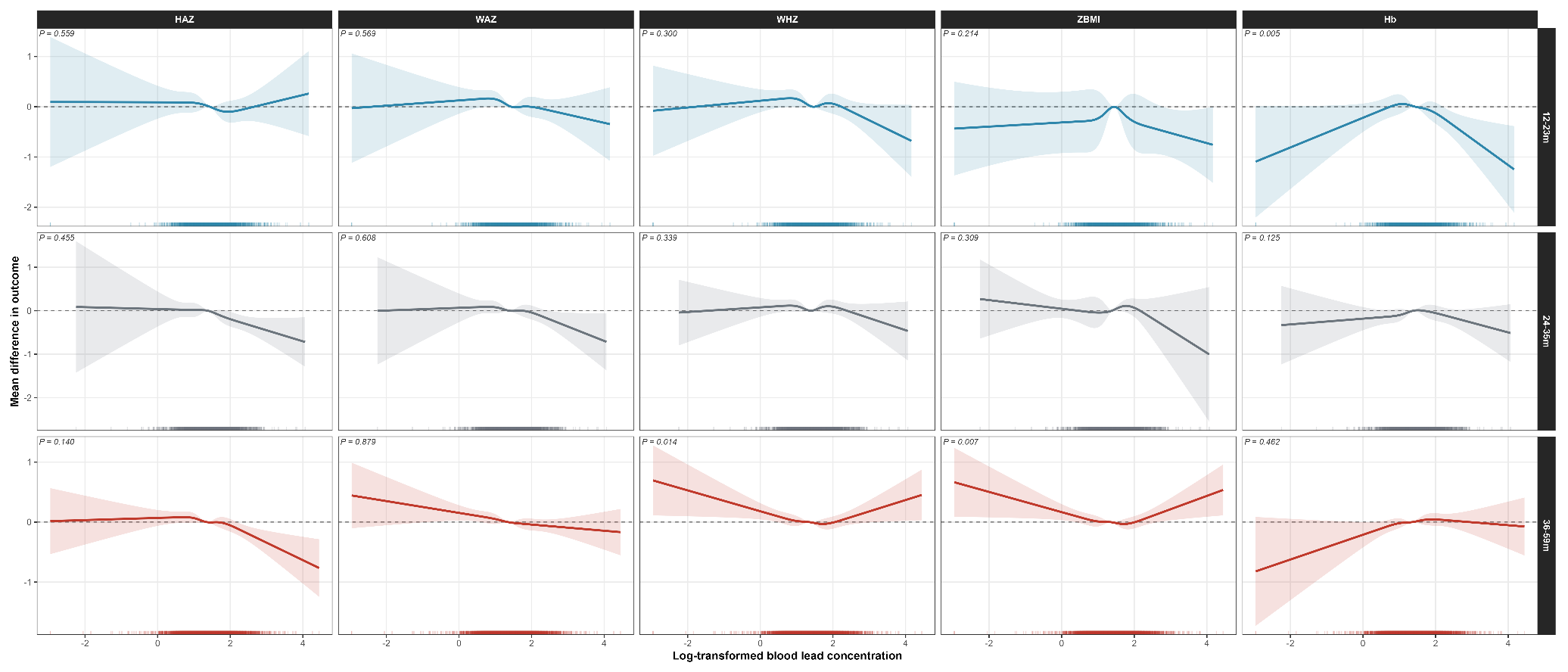

**Figure S5.** Non-linear associations of BLL (µg/dL) with child growth and haemoglobin by age group.
